# Mapping dopaminergic projections in the human brain with resting-state fMRI

**DOI:** 10.1101/2021.03.24.21254027

**Authors:** Marianne Oldehinkel, Alberto Llera, Myrthe Faber, Ismael Huertas, Jan K. Buitelaar, Bastiaan R. Bloem, Andre F. Marquand, Rick C. Helmich, Koen V. Haak, Christian F. Beckmann

## Abstract

The striatum receives dense dopaminergic projections making it a key region of the dopaminergic system. Its dysfunction has been implicated in various conditions including Parkinson’s disease and substance use disorder. However, the investigation of dopamine-specific functioning in humans is problematic as the striatum is highly interconnected and current MRI approaches are unable to differentiate between dopaminergic and other projections. Here, we demonstrate that “connectopic mapping” –a novel approach for characterizing fine-grained and overlapping modes of functional connectivity– can be used to map dopaminergic projections and as such, enables the investigation of dopamine-related functioning both in health and disease.

We applied connectopic mapping to resting-state functional MRI data of the Human Connectome Project (population cohort; *N*=839) and selected the second-order striatal connectivity mode for further analyses. We first validated its specificity to dopaminergic projections by demonstrating a very high spatial correlation (*r*=0.884) with dopamine transporter availability –a marker of dopaminergic projections– derived from DaT-SPECT scans of 209 healthy control subjects of the Parkinson’s Progression Markers Initiative. Next, we obtained the subject-specific second-order modes from 20 controls, and 39 Parkinson’s disease patients scanned under placebo and under dopamine replacement therapy (L-DOPA), and show that our proposed marker of dopamine function indeed tracks Parkinson’s disease diagnosis, symptom severity and sensitivity to L-DOPA. Finally, across 30 daily alcohol users and 38 daily smokers of the Human Connectome Project, we establish strong associations between the second-order striatal connectivity mode and self-reported weekly use of alcohol and nicotine.

Our findings provide compelling evidence that the second-order mode of functional connectivity in striatum maps onto dopaminergic projections, tracks inter-individual differences in symptom severity and L-DOPA sensitivity in Parkinson’s disease patients, and exhibits strong associations with levels of nicotine and alcohol use in a population-based cohort. We hereby provide a new biomarker for dopamine-related dysfunction in the human brain with potential clinical utility that could foster insights into the neurobiological underpinnings of various dopamine-associated conditions.

## Introduction

The brain’s dopamine system plays an important role in a wide range of behavioural and cognitive functions, including movement and reward processing.^1,2^ An integral structure of the dopamine system is the striatum, which receives dense dopaminergic projections from the substantia nigra pars compacta (SNc) and ventral tegmental area (VTA) in the midbrain.^3^ Work in experimental animals has shown that these projections organize along a gradient: dopaminergic neurons in the SNc project preferentially to dorsal caudate and putamen in dorsolateral striatum, while dopaminergic neurons in the VTA project predominantly to the nucleus accumbens (NAcc) in ventromedial striatum.^3-5^ The projections from the SNc to dorsolateral striatum comprise the nigrostriatal pathway implicated in e.g. the organization of motor planning.^1,6^ The mesolimbic pathway formed by the projections from the VTA to the NAcc has been associated with reward processing.^7,8^ In accordance with the partial neuroanatomical overlap in striatum, increasing evidence also suggests partial overlap in the function of both pathways.^9-11^ Of note, dopaminergic neurons in the VTA not only project to NAcc but also to prefrontal cortex. These cortical projections form the mesocortical pathway associated with reward-related goal-directed behaviors.^7, 8^

In humans, alterations in these dopaminergic projections have been associated with multiple neurological and psychiatric conditions.^12,13^ A well-known example is Parkinson’s disease (PD), a neurodegenerative disorder characterized by a loss of dopaminergic neurons in the SNc (part of the nigrostriatal pathway),^14^ which frequently causes asymmetric depletion of dopamine in dorsal striatum (first in putamen, later also to a lesser extent in caudate) and leads to impairments in motor- as well as a range of nonmotor functions.^15,16^ Dopaminergic dysfunction has also been implicated in substance use disorder given that addictive substances, such as stimulants, alcohol, and nicotine, increase the release of dopamine in ventral striatum (i.e., mesolimbic pathway).^17-19^

Despite the important role of the dopamine system in human brain function and its implication in disease, knowledge about this neurotransmitter system is limited and mainly based on experimental work in animals. The investigation of dopaminergic functioning in vivo in the human brain is challenging, although the nuclear imaging techniques PET and Single Photon Emission Computed Tomography (SPECT) can be used for this purpose.^20,21^ Imaging of the density of the Dopamine Transporter (DaT) using SPECT has become a popular tool to assist in the differential diagnosis of PD, as loss of dopaminergic neurons in PD is accompanied by a loss in DaT in striatum, as opposed to lookalike conditions such as dystonic tremor where the DaT signal remains intact.^22^ Tracking the loss of DaT signal over time has also been proposed as a progression biomarker for PD.^22^ Indeed, DaT reuptakes dopamine from the synaptic cleft after its release and is highly expressed in the terminals of dopaminergic neurons projecting from the midbrain to striatum.^22^ Therefore, DaT SPECT imaging can be used to image dopaminergic projections in striatum. However, the radiation exposure and costs of PET/SPECT combined with the low spatial resolution of the scan limits widespread implementation in human brain research and in clinical practice.

In this work, we hypothesize that inter-individual differences in Dopamine Transporter availability induce inter-individual variations in the synchronicity of functional activity in the brain and therefore, that dopaminergic projections in the human striatum can also be mapped using Blood-Oxygen-Level-Dependent (BOLD) functional MRI (fMRI) measured at rest. We employ a ‘connectopic mapping’ data analysis approach to disentangle striatal connectivity into multiple overlapping spatial ‘modes’ in order to dissect the complex mixture of efferent and afferent connections of the striatum to multiple cortical and subcortical systems (that map onto different neurobiological systems and associated functions).^23^ In previous work, we already showed that the dominant (zeroth-order) mode represents its basic anatomical subdivisions, while the first-order mode maps on to a ventromedial-to-dorsolateral gradient associated with goal-directed behaviour in cortex^24^ that has been described previously on the basis of tract-tracing work in non-human primates.^25^ Here, we demonstrate –by conducting a series of analyses across different datasets– that the second-order mode of gradual spatial variations in the BOLD connectivity pattern reflects Dopamine Transporter availability (DaT) in the striatum. We furthermore reveal that this mode tracks inter-individual differences in symptom severity in patients with PD, is sensitive to acute dopaminergic modulation (L-DOPA administration), and exhibits strong associations with levels of nicotine and alcohol use in a population-based cohort. Hereby we provide compelling evidence that this connectivity mode tracks inter-individual differences in dopaminergic projections and as such, offers a new biomarker for investigating dopamine-related dysfunction across various neurological and psychiatric disorders.

## Methods

### Connectopic mapping of the striatum in the HCP dataset

For our first analysis, we used resting-state fMRI data from the Human Connectome Project (HCP), which is a population-based, publicly available dataset of exceptionally high-quality.^26^ HCP participants were scanned on a customized 3 Tesla Siemens Skyra scanner (Siemens AG, Erlanger, Germany) and underwent two sessions of two 14.4 minute multi-band accelerated (TR=0.72s) resting-state fMRI scans with an isotropic spatial resolution of 2 mm. Here we included resting-state fMRI from 839 participants (aged 22-37 years; 458 females). For full details regarding the sample, data acquisition, and minimal preprocessing procedures we refer to ^26,27^. A detailed description of our preprocessing pipeline and subject inclusion criteria can be found in section 1 of the Supplementary material.

We estimated connection topographies in the striatum from the HCP resting-state fMRI data using the first session (2×14.4 minutes) for each subject. To this end, we used connectopic mapping,^23^ a novel method that enables the dominant modes of functional connectivity change within the striatum to be traced on the basis of the connectivity between each striatal voxel and the rest of the brain. We applied connectopic mapping to the left and right putamen and caudate-NAcc striatal subregions separately to increase regional specificity. When referring to connectivity modes in striatum we thus refer to the combined connectivity modes of putamen and caudate-NAcc. Masks for the striatal regions were obtained by thresholding these regions from the Harvard-Oxford atlas at 25% probability. Figure 1 summarizes the connectopic mapping procedure, which is further detailed in the Supplementary material, section 2.

**Figure 1.**
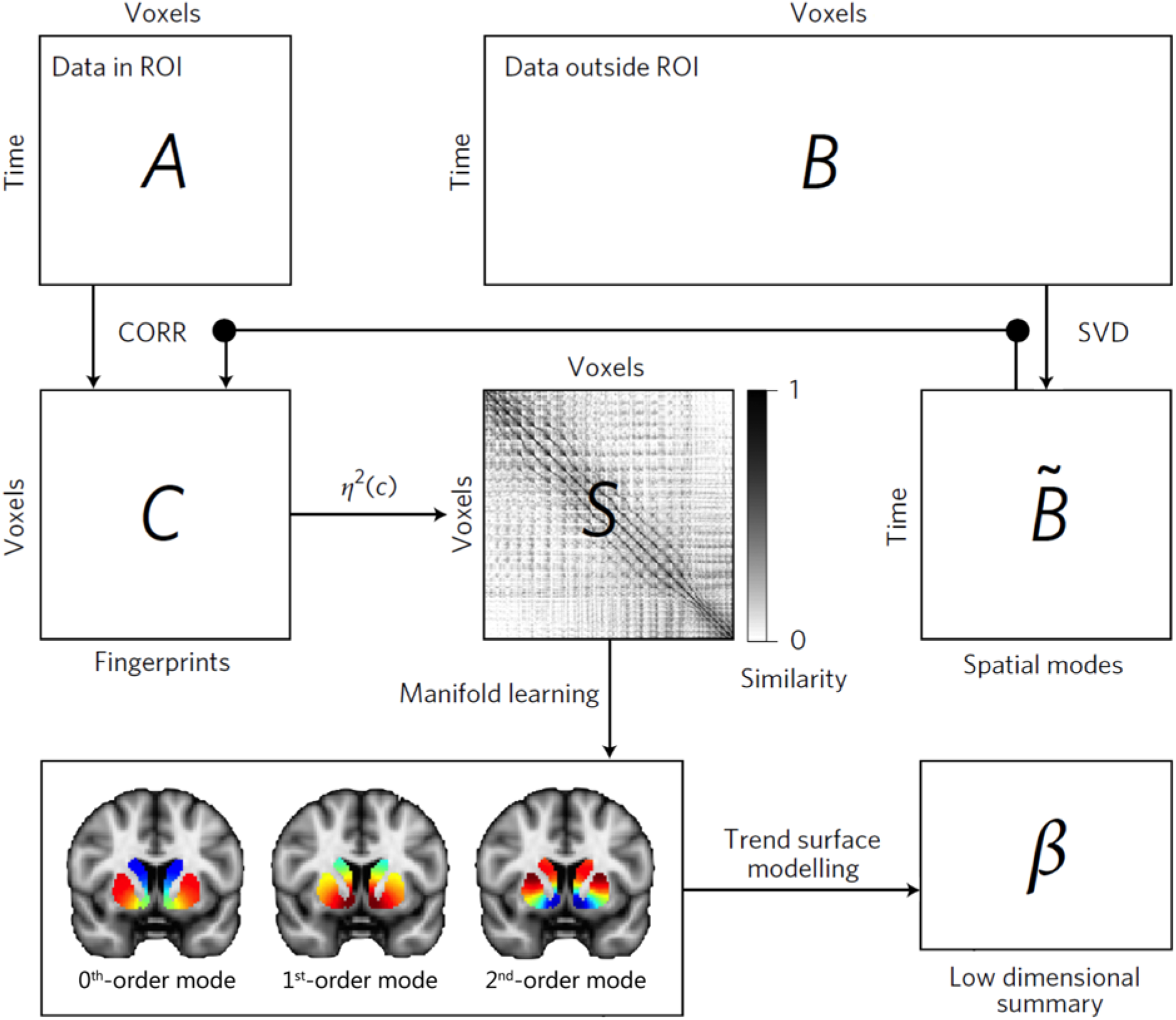
The connectopic mapping pipeline. The fMRI time-series data from a pre-defined region-of-interest (ROI), here the striatum, are rearranged into a time-by-voxels matrix *A*, as are the time serie from all voxels outside the ROI (matrix *B*). For reasons of computational tractability, the dimensionality of *B* is losslessly reduced using SVD, yielding □*B*. For every voxel within the ROI, its connectivity fingerprint is computed as the Pearson’s correlation (CORR) between the voxel-wise time-series and th SVD-transformed data, yielding matrix *C*. Then similarity between voxels is computed using the η2 coefficient, resulting in matrix *S*. Manifold learning using Laplacian eigenmaps is then applied to thi matrix, yielding a set of connection topographies or “connectivity modes” that together describe the functional organization of the striatum. Voxels that have similar colours in these connectivity modes have similar connectivity patterns with the rest of the brain. Finally, trend surface modelling is applied to summarize the connectivity modes by fitting a set of trend coefficients (β) that optimally combine a set of spatial polynomial basis functions. See Haak et al.^23^ and the supplementary material for further details.

We selected the group-average (see Figure 2) and subject-specific second-order striatal connectivity modes obtained with connectopic mapping for further analyses. The subject-specific striatal connectivity modes were highly consistent across the two fMRI sessions and showed excellent reproducibility (see Supplementary Table S1, Supplementary Figure S1). To enable statistical analysis over these connection topographies, we applied ‘trend surface modelling (TSM)’^28^ to the second-order connectivity mode of each striatal subregion for each subject, which provides an accurate representation of the topography in a small number of coefficients. A Scree plot analysis^29^ strongly favoured a polynomial of degree 2 (6 TSM coefficients) for the putamen subregion and a polynomial of degree 4 (12 TSM coefficients) for the caudate-NAcc subregion. The TSM models summarized the connectivity modes well, explaining the following *mean±s*.*d*. of the variance: left putamen: 90.5±4.16%, right putamen: 90.2±4.64%, left caudate-NAcc: 88.6±2.54%, right caudate-NAcc: 89.4±2.15%.

**Figure 2.**
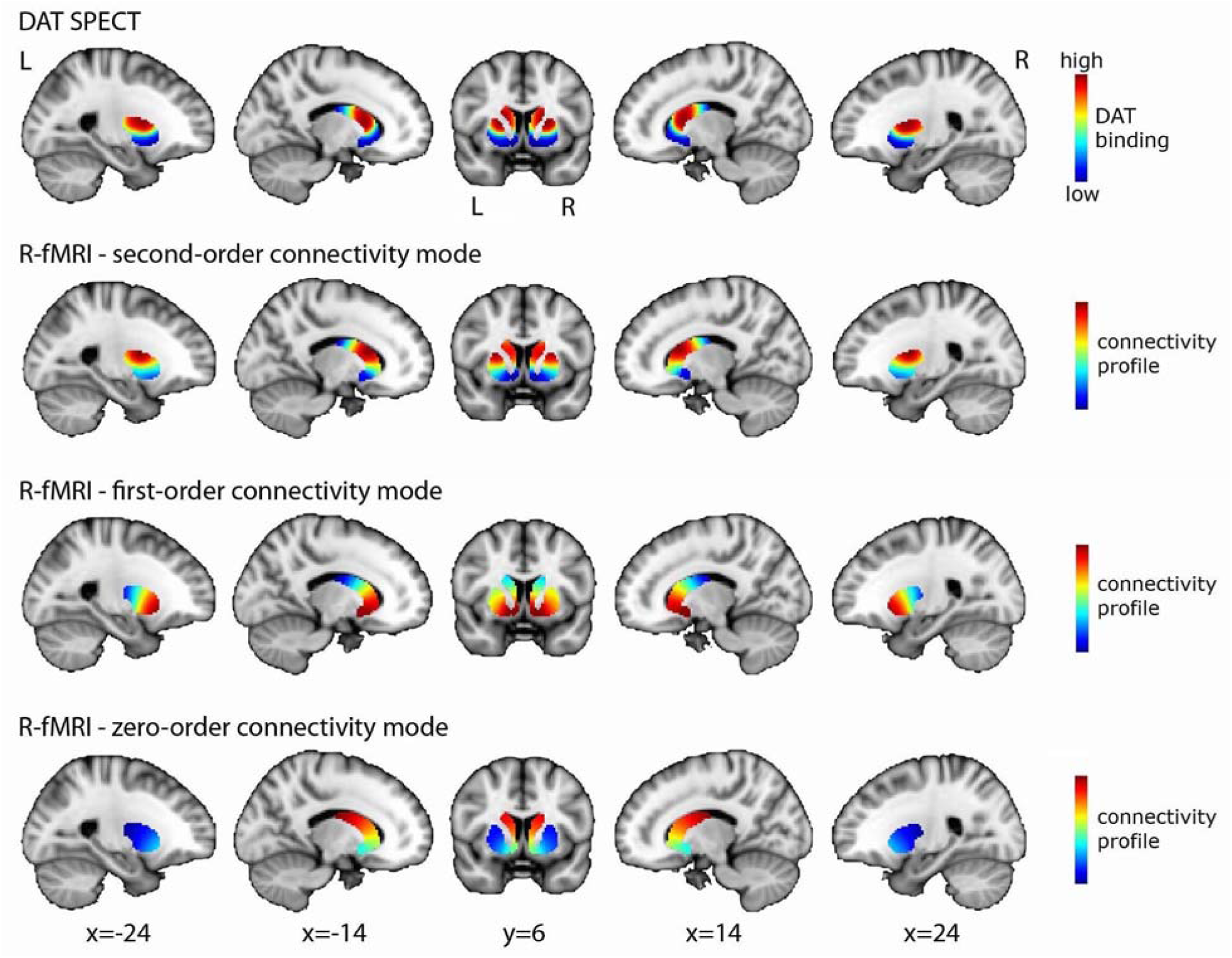
High spatial correspondence between the second-order mode of connectivity in striatum and the DaT SPECT image. The figure displays the DaT SPECT image averaged across 209 PPMI controls and the group-level connectivity modes obtained in 839 HCP subjects. The spatial correlation between the second-order mode of connectivity in striatum (modelled separately for the left and right putamen and caudate-NAcc subregions) and the DaT SPECT image is very high: *r*=0.884. To aid in visualization, the group-level modes for left and right putamen and caudate-NAcc are combined in this figure. Abbreviations: R-fMRI=resting-state fMRI, L=left, R=right.

### DaT SPECT imaging in the PPMI dataset

To determine whether the second-order striatal connectivity mode was associated with dopaminergic projections in striatum, we investigated its spatial correspondence with dopamine transporter (DaT) availability as revealed by DaT SPECT imaging. We selected DaT SPECT scans for all 209 healthy controls (aged 30-84 years; 71 females) included in the Parkinson’s Progression Markers Initiative (PPMI)^30^ database (www.ppmi-info.org/data). PPMI is a public-private partnership funded by the Michael J. Fox Foundation for Parkinson’s Research and funding partners (for up to date information, please visit www.ppmi-info.org/fundingpartners). Each participating PPMI site obtained ethical approval before study initiation and written informed consent according to the Declaration of Helsinki was obtained from all participants in the study. PPMI scans were obtained at 24 different sites and acquired with a total of seven different SPECT camera models from different manufacturers. All brain images were registered to MNI152 standard space using a linear affine transformation implemented in FSL FLIRT^31,32^ and a custom DaT SPECT template (http://www.nitrc.org/projects/spmtemplates).^33^ Typically, analysis of DaT SPECT images is limited to determining the striatal-binding ratio. This index of DaT availability is calculated by normalizing the average DaT uptake in the striatum (or a striatal sub-region) by a reference region of minimal DaT availability (e.g., cerebellum or occipital cortex). However, here we were interested in the detailed spatial profile of DaT availability across the striatum. To obtain this spatial profile, we intensity-normalized all raw DaT SPECT images^34^ so as to optimize contrast in the DaT SPECT image and take into account variability in the DaT SPECT scans across the PPMI dataset as a result of different cameras and different scan sites. Finally, we averaged across all subjects and masked the striatum to obtain the average DaT SPECT image of the striatum.

### Mapping the second-order striatal connectivity mode onto DaT availability

Next, we quantified the similarity between the second-order mode of connectivity in striatum and the average DaT SPECT image. To this end, we combined the average (i.e., group-level) second-order connectivity modes of putamen and caudate-NAcc obtained in the high-resolution HCP dataset and computed the spatial correlation of this mode with the average (i.e., group-level) DaT SPECT image of striatum obtained in the PPMI dataset. For a subsample of PPMI participants, there was next to a DaT-SPECT scan also a low-resolution resting-state fMRI scan available (130 datasets from PD patients and 14 from controls). We therefore also investigated the *within-subject* spatial correspondence between the DaT SPECT scan and the second-order connectivity mode for these subjects in the PPMI dataset. This procedure is detailed in section 4 of the Supplementary material.

### Investigating the second-order striatal mode in Parkinson’s disease

Given that PD is characterized by a loss of dopaminergic neurons,^14^ we investigated whether the second-order striatal connectivity mode was altered in PD. For this analysis, we used high-resolution resting-state fMRI data from a cohort consisting of 39 patients with PD (aged 38-81, 23 females) and 20 controls (aged 42-80, 9 females), recruited at the Centre of Expertise for Parkinson & Movement Disorders at the Radboud University Medical Center (Radboudumc) in Nijmegen and scanned at the Donders Institute in Nijmegen, The Netherlands.^35^ All patients were diagnosed with idiopathic PD (according to the UK Brain Bank criteria), and all patients had a mild to severe resting tremor besides bradykinesia. In 20 patients the motor symptoms were right-dominant, in 19 patients left-dominant. (NB: dominance here refers to the side of the body displaying the most prominent motor symptoms (including tremor)), which is believed to correspond with a dopamine depletion dominant to contralateral hemisphere in the brain). The study was approved by the local ethics committee and written informed consent according to the Declaration of Helsinki was obtained from all participants. Detailed sample characteristics can be found in Table 1.

**Table 1.**
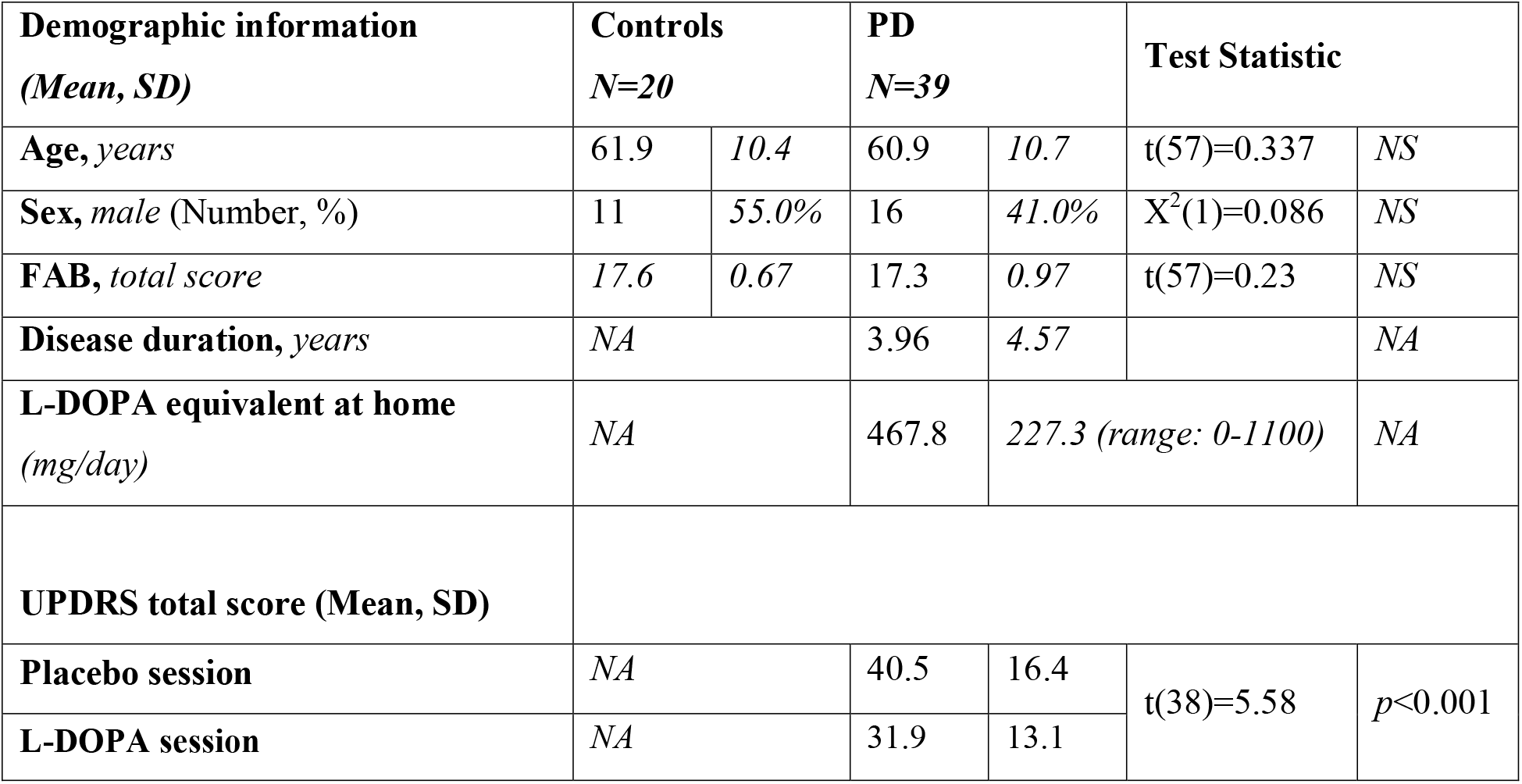
Participant characteristics. FAB=frontal assessment battery (score 0-18),^37^ UPDRS= Movement Disorder Society -Unified Parkinson’s Disease Rating Scale part III (score 0-132).^36^ For the FAB lower scores indicate worse functioning, for the UPDRS higher scores indicate worse functioning. The FAB score was evaluated OFF medication. *NS*=not significant, *NA*=not applicable, PD=Parkinson’s disease.

PD patients underwent two 10-minute resting-state fMRI sessions separated by at least a day on a 3T Siemens Magnetom Prisma^fit^ scanner. Resting-state fMRI scans were obtained with an interleaved high-resolution multiband sequence (*TR*=0.860s, voxel size=2.2mm isotropic, *TE*=34ms, flip angle=20**°**, 44 axial slices, multiband acceleration factor=4, volumes=700). During one session patients were scanned after administration of L-DOPA (a standardized dose of 200/50 mg dispersible levodopa/benserazide), a precursor of dopamine used for the treatment of PD. During the other session patients received placebo (cellulose powder). As such this dataset not only enables us to investigate the effects of clinical diagnosis but also the effects of acute dopaminergic modulation. The cellulose powder and L-DOPA/benserazide were dissolved in water and therefore undistinguishable for the participants. Patients also received 10 mg domperidone to improve gastro-intestinal absorption of levodopa and reduce side effects.

Session-order was counter-balanced and under both conditions, patients were scanned after overnight fasting in a practically defined OFF-state, i.e., more than 12h after intake of their last dose of dopaminergic medication. Symptom severity was assessed during both sessions with part III (assessment of motor function by a clinician) of the Movement Disorders Society Unified Parkinson’s Disease Rating Scale (UPDRS).^36^ In light of ethical considerations, control participants did not receive L-DOPA and placebo, they just underwent two typical resting-state fMRI sessions during which the UPDRS was not administered. We applied connectopic mapping to the pre-processed resting-state fMRI data of each session from every participant and selected the second-order connectivity mode for further analyses, using the same procedure as in the HCP dataset. Further details can be found in section 5 of the Supplementary material.

We conducted four different analyses. All these analyses were conducted separately for the left and right dominant PD groups –given that the dopamine depletion is dominant to different hemispheres in these two groups– and separately for the putamen and caudate-NAcc subregions (left+right hemisphere combined) to increase regional specificity, as PD is known to affect the putamen region of the striatum before the caudate-NAcc region. In all these analyses, we therefore corrected for multiple comparisons using a Bonferroni corrected α-level of 0.0125 (0.05/4 (2 patient groups * 2 subregions)).

In our first analysis, we compared the second-order striatal connectivity mode between the PD and control group. To this end, we conducted omnibus tests comparing the TSM coefficients modelling the second-order striatal gradient of the placebo session in the PD group with the TSM coefficients modelling the striatal gradient of the first session of the control group. More specifically, group differences in the TSM coefficients were assessed by using a likelihood ratio test in the context of a logistic regression. We report the *X*^*2*^ (likelihood test) and corresponding *p*-value of tests that revealed significant group differences. Second, since PD is a heterogeneous disease, we also conducted an analysis taking this variability into account by investigating associations between the TSM coefficients modelling the second-order striatal gradient and symptom severity in the PD group. To this end, we conducted general linear models (GLMs) that included the TSM coefficients modelling the gradient during the placebo session to predict UPDRS symptom severity scores. For all identified associations with UPDRS symptom severity, we report the *X*^*2*^ (likelihood test) and the corresponding *p*-values, and post-hoc compute Pearson correlations between UPDRS symptom severity and the individual TSM coefficients to determine which coefficients most strongly contributed to the effect.

Third, we assessed differences in the second-order striatal connectivity mode between the placebo and L-DOPA session in both PD groups. More specifically, session-differences in the TSM coefficients were assessed by using a likelihood ratio test in the context of a logistic regression. We report the *X*^*2*^ (likelihood test) and corresponding *p*-value of tests that revealed significant differences between the placebo and L-DOPA session. Finally, treatment response to L-DOPA is known to differ among patients with PD. To take this variability across patients into account, we also investigated whether the L-DOPA induced change in the second-order striatal connectivity mode was associated with L-DOPA induced changes in UPDRS symptom severity. More specifically, we calculated the difference in UPDRS symptom scores and the difference in TSM coefficients between the placebo and L-DOPA session and investigated associations within the GLM framework. For all identified associations, we post-hoc computed Pearson correlations between the change in UPDRS symptom severity and the change in individual TSM coefficients to determine which coefficients most strongly contributed to the effect.

### Investigating the second-order striatal mode in relation to tobacco and alcohol use

Given that alterations in dopaminergic functioning have also been implicated in substance use, we also investigated the association between the second-order striatal connectivity mode and tobacco use as well as alcohol use across high users within the HCP dataset. To this end, we selected HCP participants testing negative for acute drug and alcohol use but who reported to have consumed ≥3 light and/or ≥1 heavy alcoholic drinks per day over the past week (*N*=30), and participants reporting to have smoked ≥5 cigarettes every day over the past week (*N*=38). Effects of smoking and drinking were analysed separately and we again modelled the second-order striatal connectivity mode separately for left and right putamen and left and right caudate-NAcc. We conducted GLMs that included next to the amount of use over the past week (i.e., the total number of alcohol drinks or the total number of times tobacco was smoked), the TSM coefficients modelling the second-order striatal connectivity mode (i.e., the TSM coefficients modelling the putamen or caudate-NAcc). Multiple comparison correction was applied using a Bonferroni corrected α-level of 0.0125 (2 substances * 2 striatal subregions). For all identified associations with the amount of alcohol use or tobacco use, we report the *X*^*2*^ (likelihood test) and the corresponding *p*-values and post-hoc compute Pearson correlations between the amount of use and the individual TSM coefficients to determine which coefficients most strongly contributed to the association.

### Sensitivity analyses

For all the analyses described above (effects of diagnosis and L-DOPA in the PD dataset and associations with tobacco and alcohol use in the HCP dataset), we conducted post-hoc sensitivity analyses to rule out that the group differences and associations revealed by our analyses were dependent on age and sex. In addition, we investigated whether the associations with the amount of tobacco and alcohol use persisted under different usage thresholds. All these analyses are described in the Supplementary materials.

## Results

### Striatal connection topographies map onto DaT availability

Figure 2 (second row) displays the second-order connectivity mode (group-level) across striatum obtained by applying connectopic mapping to resting-state fMRI data of the HCP dataset. The group-level modes for left and right putamen and caudate-NAcc have been combined in this figure to aid in visualization and for later comparison to the DaT SPECT scan. The second-order connectivity mode comprises a gradient from the dorsal putamen and dorsal caudate (shown in red) to the ventral putamen and ventral caudate including the NAcc (shown in blue). This coding indicates that the dorsal putamen and dorsal caudate exhibit a connectivity pattern with the rest of the brain that is similar to each other but different from the ventral putamen and ventral caudate and vice versa. This striatal connectivity pattern might thus correspond with the gradient of mesolimbic and nigrostriatal dopaminergic projections to striatum (ventral versus dorsal striatum) well described by track-tracing studies in rodents and non-human primates.^3-5^ We therefore investigated it spatial correspondence to DaT-SPECT derived DaT availability in striatum, which is assumed to be an index of dopaminergic projections. As can be observed in Figure 2, the group-level second-order striatal connectivity mode indeed displays a remarkably high similarity with the group-level DaT availability in striatum, as quantified by a spatial correlation of *r*=0.884, hereby providing the first evidence for an fMRI-derived striatal connectivity marker strongly associated with dopaminergic projections into striatum. The spatial correlation of this (second-order) marker with the DaT SPECT image of the striatum was considerably higher than that of the zero-order and first-order connectivity modes (*r*=0.173 and *r*=-0.179 respectively).

In order to demonstrate that individual variations in this connectivity mode are associated with individual variations in striatal dopaminergic projections, we further aimed to replicate this mapping at the *within-subject* level in a subsample of PPMI participants with both DaT SPECT and resting-state fMRI data available. Within a smaller sample of PD patients and controls with good quality connectivity modes, we not only replicated the spatial correspondence between the connectivity mode and DaT SPECT scan at the group-level (PD group: *r*=0.714; control group: *r*=0.721) but also observed a *within-subject* spatial correlation of 0.58 (95% *CI:* [0.56,0.60]; across the four striatal subregions; see Supplementary section 4 and Supplementary Fig. 3>).

**Figure 3.**
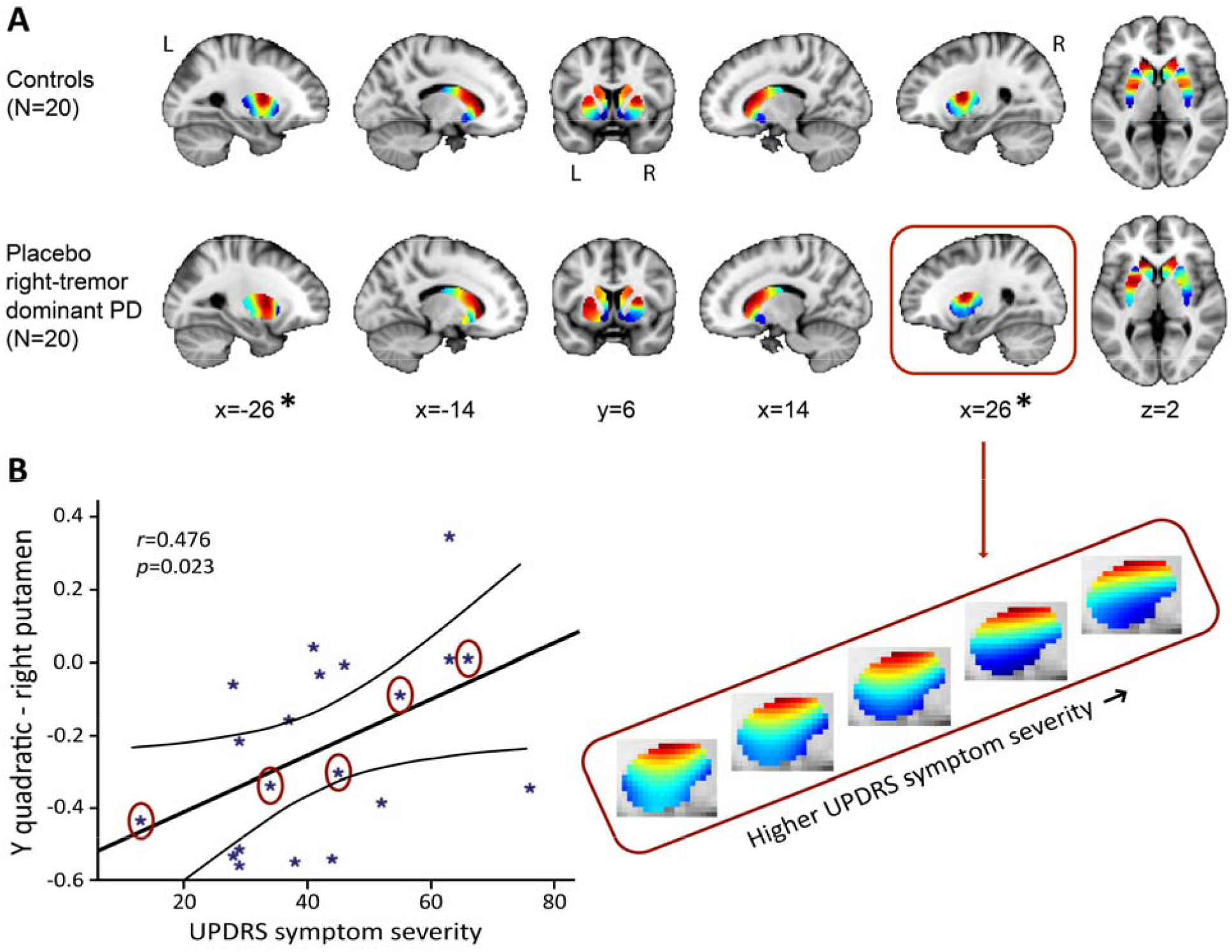
The second-order striatal connectivity mode is altered in right-dominant PD. **(A)** Significant difference between the control group and the right-dominant PD group under placebo in putamen (but not in the caudate-NAcc region; omnibus test of all TSM coefficients for putamen: *X*^*2*^=27.17, *p*=0.007). Images represent the mean connectivity modes across each of the investigated groups. The slices at MNI coordinates x=-26 and x=26 respectively show views of the striatal connectivity mode across left and right putamen were the connectivity mode is significantly different between groups (*), the slices at MNI coordinates x=-14 and x=14 respectively show views of the mode across left and right caudate-NAcc (no significant difference). **(B)** Trend-level association between total score on the motor section (part III) of the Movement Disorders Society Unified Parkinson’s Diseas Rating Scale (UPDRS) and the TSM coefficients modelling the second-order connectivity mode in putamen under placebo across patients in the right-dominant PD group (GLM omnibus test of all TSM coefficients: *X*^*2*^=22.28, *p*=0.035). Post-hoc Pearson correlations revealed that this effect was driven by th quadratic TSM coefficients modelling the striatal connectivity mode in the right putamen in the *Y* (i.e., anterior-posterior) direction (right *Y*^*2*^: *r*=0.476, *p*=0.034) and *Z* (i.e., superior-inferior) direction (right *Z*^*2*^: *r*=-0.460, *p*=0.041). The correlation between UPDRS symptom severity scores is displayed for the right Y^2^ coefficient. To visualize this association, the reconstructed second-order connectivity mode in the right putamen is shown for 5 patients with PD (data points circled) with increasing UPDRS symptom severity scores.

### Striatal connection topographies are altered in Parkinson’s disease

The PD (placebo session) vs control group analysis (first session) of the TSM coefficients modelling the second-order striatal connectivity mode, revealed a significant difference between the right-dominant PD group and the control group in the putamen region (but not in the caudate-NAcc region) of the striatum (see Figure 3A; omnibus test of all TSM coefficients: *X*^*2*^=27.17, *p*=0.007). No significant differences were observed between the left-dominant PD group and the control group. Moreover, within the right-dominant patient group, we observed a trend-level association between UPDRS symptom severity scores and the TSM coefficients modelling th putamen under placebo (GLM omnibus test of the TSM coefficients: *X*^*2*^=22.28, *p*=0.035). Post-hoc Pearson correlations revealed that this effect was driven by the quadratic TSM coefficients modelling the striatal connectivity mode in the right putamen in the *Y* (i.e., anterior-posterior) direction (right *Y*^*2*^: *r*=0.476, *p*=0.034) and *Z* (i.e., superior-inferior) direction (right *Z*^*2*^: *r*=-0.460, *p*=0.041). This association can be observed in Figure 3B, which shows an increase in blue-coded voxels in the second-order striatal connectivity mode as symptom severity increases in PD. This pattern maps very well on the observed decrease in dopaminergic projections as PD becomes clinically more severe, as reflected by higher UPDRS scores. That is, given the spatial similarity of the second-order striatal connectivity mode with the DaT SPECT scan, we can interpret the observed alteration in the connection topography as a decrease in dopaminergic projections to striatum. In Figure 3B, this is evident as an increase in blue-coded voxels and a decrease in red-coded voxels as a function of UPDRS symptom severity. Supplementary analyses showed that the observed group differences and associations with symptom severity were independent of age and sex (Supplementary Table S2).

### Striatal connection topographies are sensitive to acute dopaminergic modulation

We did not observe significant differences between the L-DOPA session and placebo session in the left-dominant and right-dominant PD groups. However, our analyses did reveal significant associations between the L-DOPA induced change in UPDRS symptom severity scores and the L-DOPA induced change in TSM coefficients in the putamen in both patient groups (GLM omnibus test of all TSM coefficients in right dominant patient group: *X*^*2*^=25.48, *p*=0.012; in left dominant patient group: *X*^*2*^=34.07, *p*=0.001). Post-hoc Pearson correlations revealed that these effects were driven by the linear TSM coefficient (Y^1^) modelling the striatal connectivity mode in putamen in the *Y* (i.e., anterior-posterior) direction (right dominant PD: left *Y*^*1*^: *r*=-0.548, *p*=0.012; left dominant PD: right *Y*^*1*^: *r*=0.345, *p*=0.15 (strongest Pearson correlation but did not reach significance)). As can be seen in Figure 4, a larger L-DOPA induced reduction in UPDRS scores is associated with a larger positive change (i.e., an increase in red-coded voxels) in the superior-anterior part of the putamen, which we hypothesize maps onto an increase in dopamine-related connectivity. Supplementary analyses demonstrated that the observed effects of L-DOPA were independent of age and sex (see Supplementary Table S2).

**Figure 4.**
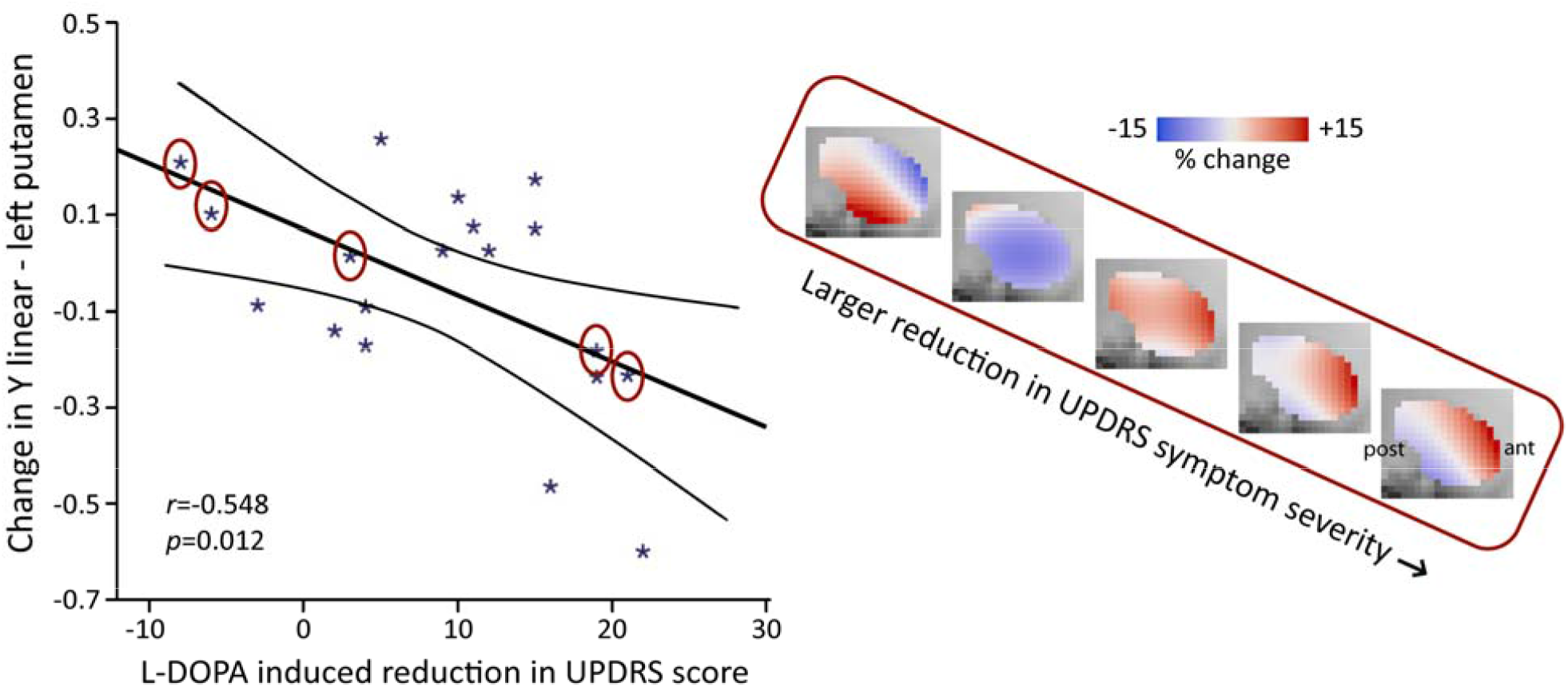
L-DOPA induced reduction in UPDRS symptom severity score (total score on motor section) is associated with the L-DOPA induced change in the second-order mode of connectivity in putamen in right dominant PD. (GLM omnibus test of all TSM coefficients modelling putamen for right tremor dominant PD: *X*^*2*^=25.48, *p*=0.012). Post-hoc Pearson correlations revealed that this effect was driven by the linear TSM coefficient modelling the striatal connectivity mode in the left putamen in the *Y* direction (left *Y*^*1*^: *r*=-0.548, *p*=0.012). To visualize this association, the difference in the reconstructed second-order connectivity modes between the placebo and L-DOPA session is shown for the left putamen (at slice X=-24) for 5 patients with PD (data points circled). Red-coded voxels are hypothesized to map onto an increase in dopaminergic connectivity, blue-coded voxels onto a decrease. A significant effect was also observed for the left-dominant PD group (GLM omnibus test of all TSM coefficients: *X*^*2*^=34.07, *p*=0.001), but as post-hoc Pearson correlations did not reveal significant associations with one of the individual TSM coefficients in this group, this association is not shown. Abbreviations: ant=anterior, post=posterior putamen.

### Striatal connection topographies are associated with the amount of substance use

In smokers of the HCP dataset, we observed a significant association with the total number of cigarettes smoked over the past week for the TSM coefficients modelling the second-order connectivity mode in the caudate-NAcc region (*X*^*2*^=49.55, *p*=0.002), but not for the putamen region. Subsequent computation of the Pearson correlations between the individual TSM coefficients and the amount of use revealed that this association was driven by multiple TSM coefficients in both the left and right caudate-NAcc (left *X*^*3*^: *r*=-0.409, *p*=0.011, right *Z*^*1*^: *r*=0.408, *p*=0.011, right *Y*^*3*^: *r*=0.367, *p*=0.024, right *Z*^*3*^: *r*=-0.451, *p*=0.005, right *Y*^*4*^: *r*=0.362, *p*=0.026; see Supplementary Figure S4). As can be observed in Figure 5 (top panel), alterations in the second-order striatal connectivity mode are subtle but consist of an increase in inferior blue-coded voxels in the caudate-NAcc as tobacco use in this population cohort increases.

**Figure 5.**
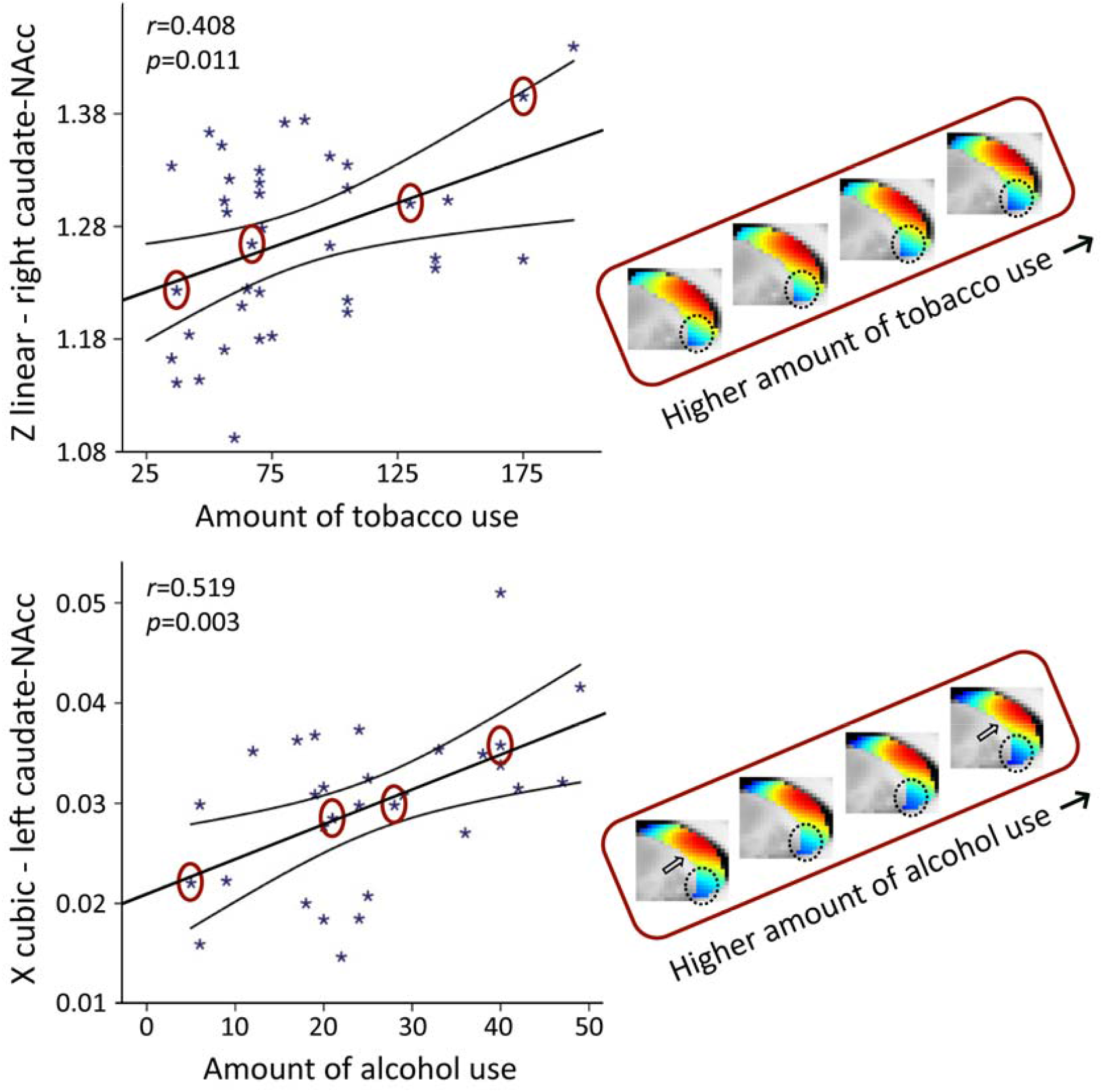
The second-order mode of connectivity in striatum is associated with the amount of tobacco use (top) and alcohol use (bottom). Strong associations were observed between the TSM coefficients modelling the connectivity mode in the caudate-NAcc region and the total amount of tobacco use as well as alcohol use over the past week (GLM omnibus test tobacco use: *X*^*2*^=49.55, *p*=0.002; alcohol use: *X*^*2*^=64.45, *p*<0.001). To visualize these relationships, Pearson correlations between one of the significant TSM coefficients and the amount of use are shown as well as the reconstructed second-order connectivity mode in the right caudate-NAcc (at slice X=14) for 4 tobacco users and in the left caudate-NAcc (at slice X=-14) for 4 alcohol users with increasing of amounts of use (data points circled). Circles and arrows indicate where in the connectivity mode tobacco and alcohol use-related changes can be observed. Correlation plots for the other TSM coefficients can be found in Supplementary Figures S4 and S5.

In the heavy drinkers, we also observed a strong association with the total number of alcoholic drinks consumed over the past week for the TSM coefficients modelling the connectivity mode in the caudate-NAcc region (*X*^*2*^=64.45, *p*<0.001), but again not for the putamen region. Subsequent computation of the Pearson correlations between the individual TSM coefficients and the amount of use, revealed that this association was driven by multiple TSM coefficients in both the left and right caudate-NAcc (left *X*^*1*^: *r*=-0.378, *p*=0.039, left *Y*^*1*^: r=0.399, p=0.029, left *Y*^*2*^: *r*=0.488, *p*=0.006, left *Z*^*2*^: *r*=-0.386, *p*=0.035, left *X*^*3*^: *r*=0.519, *p*=0.003, left *Y*^*4*^: *r*=-0.417, *p*=0.022, right *Z*^*2*^: *r*=-0.418, *p*=0.022, right *Z*^*4*^: *r*=0.488, *p*=0.006; see Supplementary Fig. S5). Similar to tobacco use, Figure 5 (bottom panel) shows that higher levels of alcohol use are accompanied by a subtle increase in blue-coded voxels as well as a decrease in red-coded voxels in the caudate-NAcc region of the second-order connectivity mode. We argue that these subtle increases in blue-coded voxels (and decrease in red-coded voxels) in high nicotine and alcohol users map onto decreases in dopaminergic connectivity, which corresponds with reported reductions in dopamine release in striatum in patients with nicotine and alcohol dependence.^19,38^ Supplementary analyses revealed that the associations with the amount of tobacco and alcohol use persisted under different usage thresholds and were independent of age and sex (see Supplementary Tables S2-S4).

## Discussion

In this work, we provide evidence for a resting-state fMRI-derived biomarker of dopamine function in the striatum in man. Specifically, we demonstrated that one particular mode of functional connectivity in the striatum showed a remarkably high spatial correspondence to dopamine transporter availability (DaT), a marker of dopaminergic projections derived from DaT SPECT imaging. This observation generated multiple hypotheses that we validated using both data from PD patients and healthy controls. We showed that this second-order striatal connectivity mode is associated with symptom severity and sensitive to acute dopaminergic modulation by L-DOPA in persons with PD, a disorder characterized by a degeneration of dopaminergic neurons projecting to striatum.^14-16^ We also demonstrated that this mode is associated with the amount of tobacco and alcohol use, both of which have been related to alterations in dopaminergic signalling.^17-19^ As such, our results provide compelling evidence that the second-order mode of functional connectivity in striatum maps onto dopaminergic projections and can be used as a non-invasive biomarker for investigating dopaminergic (dys)function in PD and substance use. Formal quantification of test-retest reliability suggests that this gradient approach has very high measurement consistency and therefore lends itself for further investigation into the clinical utility across the various neurological and psychiatric disorders associated with dopaminergic functioning.

By applying connectopic mapping, we shift away from the vast majority of resting-state fMRI studies that employ hard parcellations, which only allow investigating the average functional connectivity signal in one or more regions of interest, thereby ignoring both the topographic organization of and functional multiplicity in the brain. In contrast, our approach not only enables characterization of smooth, gradual changes in functional connectivity, but it also enables the detection of multiple, overlapping modes of functional connectivity in a region that might exist at the same time.^39^ The work presented here signifies the importance of both features, by not only showing that the second-order striatal connectivity mode comprises a smooth gradient from the dorsal putamen and dorsal caudate to the ventral putamen and ventral caudate including the NAcc, but also that this second-order mode –but not the zeroth-order or first-order mode– maps onto DaT availability. In doing so, we are the first to demonstrate a direct mapping between a functional connectivity-derived marker and dopaminergic projections.

DaT is highly expressed in the terminals of dopaminergic neurons projecting from the midbrain to striatum.^22^ The high spatial correlation (*r*=0.884) between the group-average second-order connectivity mode in the HCP dataset and the group-average DaT SPECT image in the PPMI dataset as well as the significant within-subject correlation between the connectivity mode and DaT SPECT scan (*r*=0.58) in PPMI subjects, therefore suggests that this connectivity mode maps onto these dopaminergic projections to striatum. Animal work has shown that dopaminergic projections form a gradient with nigrostriatal neurons from SNc projecting predominantly to the dorsolateral striatum (putamen and caudate) and mesolimbic neurons from the VTA projecting predominantly to the ventromedial striatum (NAcc),^3-5^ representing a functional connectivity gradient formed by the SNc projections to the dorsolateral (putamen/caudate) and VTA projections to the ventromedial striatum (NAcc). Studies demonstrating that the average striatal DaT binding as obtained by DaT SPECT or PET imaging is highly correlated with averaged post-mortem SN cell counts in humans are in support of this view.^40-42^ However, to our knowledge the relationship between DaT SPECT/PET and VTA cell counts in humans has not been investigated, and future work will thus be necessary to determine the exact relationship between this striatal connectivity mode, DaT availability assessed by DaT-SPECT, and dopaminergic projections.

While we were able to replicate the spatial correlation between the second-order connectivity mode and the DaT SPECT scan at the *within-subject* level, this spatial correlation (*r*=0.58) is not as high as the spatial correlations observed at the group level (i.e., *r*=0.721 and *r*=0.714 for PPMI controls and PD patients respectively, and *r*=0.884 between the DaT SPECT scan in PPMI controls and the connectivity mode in HCP participants). This is not surprising given the relatively low temporal resolution of the resting-state fMRI scan of the PPMI dataset (TR=2400ms, 260 volumes). While this resolution is sufficient for typical resting-state fMRI analyses at the group-level, the precise delineation of the very fine-grained and overlapping connectivity modes using connectopic mapping at the single-subject level calls for high spatial and temporal resolution data.^23^ However, to our knowledge, there is currently no dataset publicly available that includes both a high-resolution resting-state fMRI scan and a DaT-SPECT scans from the same participants.

Nevertheless, adding to its association with dopaminergic projections are the alterations of this connectivity mode observed in PD. This disorder is characterised by a loss of nigrostriatal dopaminergic neurons projecting from SNc to the striatum, which is most prominent in the putamen.^14^ Corresponding to the pathology of the condition, we observed a significant difference of this connectivity mode between right dominant PD patients and control participants in the putamen. Moreover, within this patient group the second-order striatal connectivity mode was also sensitive to inter-subject variability as revealed by the association with symptom severity. This association is visualized in Figure 3B, which shows that portions of the gradient that map on low DaT availability (blue) increase as symptom severity in PD increases. We argue that the second-order striatal connectivity mode hereby follows the expected pattern of a reduction in dopaminergic projections to the putamen (as indexed by decreased DaT availability) as symptom severity increases in PD. It should be noted though that we did not find a significant difference between controls and the left dominant PD group. Since there is no evidence that different mechanisms underlie left-dominant and right-dominant PD (apart from the difference in the most-affected hemisphere), this might be a power issue that requires further investigation.

Not only was the second-order mode of connectivity in striatum sensitive to variability in symptom severity in a clinical cohort, but also to behavioural variability associated with self-reported alcohol and tobacco use in a healthy, non-clinical population. Substance use has frequently been associated with alterations in dopamine release in the ventromedial striatum (NAcc), part of the mesolimbic dopaminergic pathway.^17-19^ Corresponding with these findings, we observed significant associations with the amount of alcohol and tobacco use over the past week in the caudate-NAcc region, but not in the putamen region of the second-order striatal connectivity mode. As such, not only are these alterations in PD and high alcohol and tobacco users of the HCP dataset consistent with the hypothesis that the second-order striatal connectivity mode reflects dopaminergic projections, the alterations are also specific to the hypothesized striatal subregions and dopaminergic pathways. That is, we found that PD –a disorder characterized by death of nigrostriatal dopaminergic neurons leading to motor impairments– was associated with connectivity alterations in the putamen, which is a key region of the nigrostriatal pathway that has predominantly been implicated in motor function.^1, 6^ On the other hand, tobacco and alcohol use were associated with connectivity alterations in the caudate-NAcc region, which is part of the mesolimbic pathway that has repeatedly been implicated in reward processing and substance use.^7, 8^

Finally, we observed that the change in the second-order mode of connectivity in striatum induced by L-DOPA administration was associated with the change in symptom severity in PD patients. L-DOPA is used as a drug for the treatment of PD, yet not all patients are equally responsive to L-DOPA treatment. When L-DOPA crosses the blood-brain barrier it is converted into dopamine and is assumed to increase dopaminergic signalling.^43^ However, there are differences between PD patients in treatment response, which can be explained by a variety of factors, including differences in the level of systemic L-DOPA uptake from the gut.^44^ Our finding thus indicates that the second-order striatal connectivity mode is differentially sensitive to acute dopaminergic modulation across patients with PD and that the amount of this change is associated with the amount of change in symptom severity. This adds to our hypothesis that this mode is associated with dopamine-related functional connectivity and furthermore indicates that studying the dopaminergic system by applying connectopic mapping to resting-state fMRI offers advantages over PET and SPECT scans: PET and SPECT are not only invasive and limited by their low spatial resolution but also depend on indirect measures of dopaminergic signalling such as availability of dopamine transporters and receptors and are therefore not very sensitive to acute, temporal alterations in dopaminergic signalling. In contrast, here we show that connectopic mapping does allow for the investigation of both fine-grained spatial and short-term temporal changes in dopamine-related functional connectivity.

In conclusion, our results provide compelling evidence that the second-order mode of resting-state functional connectivity in striatum is associated with dopaminergic projections and can be used as a non-invasive biomarker for investigating dopaminergic (dys)function. This has wide-ranging clinical and scientific applications across disorders associated with dopaminergic functioning. For example, in the diagnostic work-up of movement disorders where DaT-SPECT is currently used to distinguish between PD and essential tremor or dystonic tremor, the resting-state fMRI derived second-order connectivity mode might be used instead. The correlation with symptom severity suggests that the resting-state fMRI might also be used as a progression biomarker, for example to track differences in rate of progression in future intervention studies of new experimental medications aimed at modifying the course of PD. Our results furthermore suggest that this striatal connectivity mode is associated with functions of both the nigrostriatal and mesolimbic pathway, and that it might be possible to differentiate between the two dopaminergic pathways by considering in which striatal subregion that gradient is altered: connectivity alterations seem to occur in putamen for functions associated with the nigrostriatal pathway and in ventral caudate/NAcc for functions associated with the mesolimbic pathway. However, the exact mapping of this striatal connectivity mode on both pathways as well as its relation with the first-order, ventromedial-to-dorsolateral striatal gradient, which we previously linked to goal-directed behaviours, is subject for further investigation.

## Data Availability

We made use of publicly available data from the HCP dataset (S1200 release) and from the PPMI dataset (see https://www.humanconnectome.org/study/hcp-young-adult/document/1200-subjects-data-release and https://www.ppmi-info.org/access-data-specimens/download-data). The subject identifiers from the HCP and PPMI datasets used in our analyses can be found in Supplementary Tables S5-S7. Please note that the subject identifiers from the subset of HCP subjects included in the nicotine-use and alcohol-use analyses cannot be provided, since these are based on the HCP restricted data. The code used for the connectopic mapping procedure is available at the following Github repository: https://github.com/koenhaak/congrads. The remaining data and analysis code are available from the corresponding author upon reasonable request.

## Funding and Acknowledgements

This work was supported by the Netherlands Organization for Scientific Research Vidi Grant No. 864-12-004 (to CFB), Vici Grant No. 17854 (to CFB), Vidi Grant No. 016.156.415 (to AM), Veni Grant No. 016.171.068 (to KVH), Veni Grant No. 91617077 (to RH) and NWO-CAS Grant No. 012-200-013 (to CFB, providing funds to MF). MO was supported by ZonMW Rubicon Grant No. 452172019. RH was also funded by a grant of the Dutch Brain Foundation grant F2013(10–15). The Centre of Expertise for Parkinson & Movement Disorders of the Radboud University Medical Centre was supported by a centre of excellence grant of the Parkinson’s Foundation.

We further made use of HCP data that were provided by the Human Connectome Project, WU-Minn Consortium (Principal Investigators: David Van Essen and Kamil Ugurbil; 1U54MH091657) funded by the 16 NIH Institutes and Centres that support the NIH Blueprint for Neuroscience Research; and by the McDonnell Centre for Systems Neuroscience at Washington University. We also used data from the Parkinson’s Progression Markers Initiative (PPMI) database (www.ppmi-info.org/data). PPMI—a public-private partnership—is funded by the Michael J. Fox Foundation for Parkinson’s Research funding partners Abbvie, Avid Radiopharmaceuticals, Biogen Idec, BioLegend, Bristol-Myers Squibb, Eli Lilly & Co., F. Hoffman-La Roche, Ltd., GE Healthcare, Genentech, GlaxoSmithKline, Lundbeck, Merck, MesoScale Discovery, Piramal, Pfizer, Sanofi Genzyme, Servier, Takeda, Teva, and UCB.

## Competing interests

CFB is director and shareholder in SBGneuro Ltd. All other authors report no competing interests.

## Notes

### Author Declarations

Medical-ethical committee at the Radboud University Medical Center and local ethical committees from centers involved in the collection of the publicly available data from the Human Connectome Project (HCP) and Parkinson's Progression Markers Initiative (PPMI)

## References

1. Joshua M, Adler A, Bergman H. The dynamics of dopamine in control of motor behavior. Current opinion in neurobiology. 2009;19(6):615–620.

2. Ruhé HG, Mason NS, Schene AH. Mood is indirectly related to serotonin, norepinephrine and dopamine levels in humans: a meta-analysis of monoamine depletion studies. Molecular psychiatry. 2007;12(4):331.

3. Steiner H, Tseng KY. Handbook of basal ganglia structure and function. Vol 24: Academic Press; 2016.

4. Haber SN. The place of dopamine in the cortico-basal ganglia circuit. Neuroscience. 2014;282:248–257.

5. Björklund A, Dunnett SB. Dopamine neuron systems in the brain: an update. Trends in neurosciences. 2007;30(5):194–202.

6. Faure A, Haberland U, Condé F, El Massioui N. Lesion to the nigrostriatal dopamine system disrupts stimulus-response habit formation. Journal of Neuroscience. 2005;25(11):2771–2780.

7. Schultz W. Updating dopamine reward signals. Current opinion in neurobiology. 2013;23(2):229–238.

8. Wise RA. Dopamine, learning and motivation. Nature reviews neuroscience. 2004;5(6):483.

9. Haber SN, Fudge JL, McFarland NR. Striatonigrostriatal pathways in primates form an ascending spiral from the shell to the dorsolateral striatum. Journal of Neuroscience. 2000;20(6):2369–2382.

10. Everitt BJ, Robbins TW. Neural systems of reinforcement for drug addiction: from actions to habits to compulsion. Nature neuroscience. 2005;8(11):1481.

11. Wise RA. Roles for nigrostriatal—not just mesocorticolimbic—dopamine in reward and addiction. Trends in neurosciences. 2009;32(10):517–524.

12. DeLong MR, Wichmann T. Circuits and circuit disorders of the basal ganglia. Archives of neurology. 2007;64(1):20–24.

13. Money KM, Stanwood GD. Developmental origins of brain disorders: roles for dopamine. Frontiers in cellular neuroscience. 2013;7:260.

14. Fearnley JM, Lees AJ. Ageing and Parkinson’s disease: substantia nigra regional selectivity. Brain. 1991;114(5):2283–2301.

15. Brooks DJ, Piccini P. Imaging in Parkinson’s disease: the role of monoamines in behavior. Biological psychiatry. 2006;59(10):908–918.

16. Hornykiewicz O. Basic research on dopamine in Parkinson’s disease and the discovery of the nigrostriatal dopamine pathway: the view of an eyewitness. Neurodegenerative Diseases. 2008;5(3-4):114–117.

17. Laruelle M, Abi-Dargham A, van Dyck CH, et al. SPECT imaging of striatal dopamine release after amphetamine challenge. Journal of Nuclear Medicine. 1995;36(7):1182–1190.

18. Barrett SP, Boileau I, Okker J, Pihl RO, Dagher A. The hedonic response to cigarette smoking is proportional to dopamine release in the human striatum as measured by positron emission tomography and [11C] raclopride. Synapse. 2004;54(2):65–71.

19. Nutt DJ, Lingford-Hughes A, Erritzoe D, Stokes PR. The dopamine theory of addiction: 40 years of highs and lows. Nature Reviews Neuroscience. 2015;16(5):305.

20. Blake P, Johnson B, VanMeter JW. Positron emission tomography (PET) and single photon emission computed tomography (SPECT): clinical applications. Journal of neuro-ophthalmology. 2003;23(1):34–41.

21. Volkow ND, Fowler JS, Gatley SJ, Logan J. PET evaluation of the dopamine system of the human brain. The Journal of nuclear medicine. 1996;37(7):1242.

22. Brooks DJ. Molecular imaging of dopamine transporters. Ageing research reviews. 2016;30:114–121.

23. Haak KV, Marquand AF, Beckmann CF. Connectopic mapping with resting-state fMRI. NeuroImage. 2018/04/15/ 2018;170:83–94.

24. Marquand AF, Haak KV, Beckmann CF. Functional corticostriatal connection topographies predict goal-directed behaviour in humans. Nature human behaviour. 2017;1:s41562-41017-40146.

25. Haber SN, Knutson B. The reward circuit: linking primate anatomy and human imaging. Neuropsychopharmacology. 2010;35(1):4–26.

26. Van Essen DC, Smith SM, Barch DM, et al. The WU-Minn human connectome project: an overview. Neuroimage. 2013;80:62–79.

27. Glasser MF, Sotiropoulos SN, Wilson JA, et al. The minimal preprocessing pipelines for the Human Connectome Project. Neuroimage. 2013;80:105–124.

28. Gelfand AE, Diggle P, Guttorp P, Fuentes M. Handbook of spatial statistics: CRC press; 2010.

29. Cattell RB. The scree test for the number of factors. Multivariate behavioral research. 1966;1(2):245–276.

30. Marek K, Jennings D, Lasch S, et al. The parkinson progression marker initiative (PPMI). Progress in neurobiology. 2011;95(4):629–635.

31. Jenkinson M, Smith S. A global optimisation method for robust affine registration of brain images. Medical image analysis. 2001;5(2):143–156.

32. Jenkinson M, Bannister P, Brady M, Smith S. Improved optimization for the robust and accurate linear registration and motion correction of brain images. Neuroimage. 2002;17(2):825–841.

33. Garcia-Gomez F, García-Solís D, Luis-Simón F, et al. Elaboration of the SPM template for the standardization of SPECT images with 123I-Ioflupane. Revista Española de Medicina Nuclear e Imagen Molecular (English Edition). 2013;32(6):350–356.

34. Llera A, Huertas I, Mir P, Beckmann CF. Quantitative Intensity Harmonization of Dopamine Transporter SPECT Images Using Gamma Mixture Models. Molecular Imaging and Biology. 2018:1–9.

35. Dirkx MF, Zach H, van Nuland A, Bloem BR, Toni I, Helmich RC. Cerebral differences between dopamine-resistant and dopamine-responsive Parkinson’s tremor. Brain. 2019;142(10):3144–3157.

36. Goetz CG, Tilley BC, Shaftman SR, et al. Movement Disorder SocietylJsponsored revision of the Unified Parkinson’s Disease Rating Scale (MDSlJUPDRS): scale presentation and clinimetric testing results. Movement disorders: official journal of the Movement Disorder Society. 2008;23(15):2129–2170.

37. Dubois B, Slachevsky A, Litvan I, Pillon B. The FAB: a frontal assessment battery at bedside. Neurology. 2000;55(11):1621–1626.

38. Balfour DJ. The role of mesoaccumbens dopamine in nicotine dependence. The Neuropharmacology of Nicotine Dependence: Springer; 2015:55–98.

39. Haak KV, Beckmann CF. Understanding brain organisation in the face of functional heterogeneity and functional multiplicity. NeuroImage. 2020:117061.

40. Snow B, Tooyama I, McGeer E, et al. Human positron emission tomographic [18F] fluorodopa studies correlate with dopamine cell counts and levels. Annals of Neurology: Official Journal of the American Neurological Association and the Child Neurology Society. 1993;34(3):324–330.

41. Colloby SJ, McParland S, O’brien JT, Attems J. Neuropathological correlates of dopaminergic imaging in Alzheimer’s disease and Lewy body dementias. Brain. 2012;135(9):2798–2808.

42. Kraemmer J, Kovacs GG, PerjulJDumbrava L, Pirker S, TraublJWeidinger T, Pirker W. Correlation of striatal dopamine transporter imaging with post mortem substantia nigra cell counts. Movement Disorders. 2014;29(14):1767–1773.

43. LeWitt PA. Levodopa for the treatment of Parkinson’s disease. New England Journal of Medicine. 2008;359(23):2468–2476.

44. Nonnekes J, Timmer MH, de Vries NM, Rascol O, Helmich RC, Bloem BR. Unmasking levodopa resistance in Parkinson’s disease. Movement Disorders. 2016;31(11):1602–1609.

