## Supplementary_Materials for "Mapping dopaminergic projections in the human brain with resting-state fMRI"

### 1. Resting-state fMRI data of the Human Connectome Project dataset

We used resting-state fMRI data from the Human Connectome Project (HCP), an exceptionally high-quality, publicly available neuroimaging dataset.[1] HCP participants were scanned on a customized 3 Tesla Siemens Skyra scanner (Siemens AG, Erlanger, Germany) and underwent two sessions of two 14.4 minute multi-band accelerated (TR=0.72s) resting-state fMRI scans with an isotropic spatial resolution of 2mm. Here, we included participants from the S1200 release who completed at least one resting-state fMRI session (2x14.4 minutes) and for whom data was reconstructed with the r227 reconstruction algorithm. (The reconstruction algorithm was upgraded in late April 2013 from the original 177 ICE version to the 227 upgraded ICE version. As the reconstruction version has been shown to make a notable signature on the data that can make a large difference in fMRI data analysis (for details see https://wiki.humanconnectome.org/display/PublicData/Ramifications+of+Image+Reconstruction+Version+Differences), we only included participants with r227 reconstructions). This resulted in the inclusion of 839 participants (aged 22-37 years; 458 females). Resting-state fMRI data were preprocessed according to the HCP minimal processing pipeline[2] which included corrections for spatial distortions and head motion, registration to the T1w structural image, resampling to 2mm MNI152 space, global intensity normalization and high-pass filtering with a cut-off at 2000s. The data were subsequently denoised using ICA-FIX –an advanced independent component analysis-based artefact removal procedure[3]–, and smoothed with a 6mm kernel.

### 2. Connectopic mapping of the striatum in the Human Connectome Project dataset

We estimated connection topographies from the HCP resting-state fMRI data using the first session (2x14.4 minutes) for each subject. To this end, we used connectopic mapping,[4] a novel method that enables the dominant modes of functional connectivity change within the striatum to be traced on the basis of the connectivity between each striatal voxel and the rest of the brain (see Figure 1). In previous work we showed that the dominant mode (zeroth-order mode) of connectivity in the striatum obtained with connectopic mapping represented its anatomical subdivision into putamen, caudate and NAcc. Since higher-order modes are restricted by lower-order modes, we decided to take the anatomical subdivision in the striatum into account by applying connectopic mapping in the current work to the left and right putamen and caudate-NAcc striatal subregions separately, thereby also increasing regional specificity. When referring to the second-order mode of connectivity in striatum we thus refer to the combination of the second-order connectivity modes of putamen and caudate-NAcc. We did not apply connectopic mapping to the NAcc and caudate separately as the left NAcc and right NAcc only include 136 voxels and 127 voxels respectively. We expect that this very small region is too homogenous in terms of connectivity with cortex to estimate reliable overlapping connectivity modes. Masks for the striatal regions were obtained by thresholding the respective regions from the Harvard-Oxford atlas at 25% probability.

In brief, we rearranged the fMRI time-series data from each striatal subregion and all grey-matter voxels outside the striatum into two time-by-voxels matrices. Since the latter is relatively large, we reduced its dimensionality using a lossless singular value decomposition (SVD). We then computed the correlation between the voxel-wise striatal time-series data and the SVD-transformed data from outside the striatum, and subsequently used the $\eta^2$ coefficient to quantify the similarities among the voxel-wise fingerprints.[4] Next, we applied the Laplacian eigenmaps non-linear manifold learning algorithm[5] to the acquired similarity matrix, which resulted in a series of vectors representing the dominant modes of functional connectivity change. Note that this can be done at the group level by using the average of the individual similarity matrices or individually for each subject (as used for statistical analysis). We selected the second-order striatal connectivity mode (both the average and subject-specific modes) for further analyses.

Finally, to enable statistical analysis over these connection topographies, we fitted spatial statistical models to the second-order connectivity mode of each striatal subregion to provide an accurate representation of the topography in a small number of coefficients. For this, we use a 'trend surface modelling' (TSM) approach,[6] which involves fitting a set of polynomial basis functions defined by the coordinates of each striatal location to predict each individual subject's connection topography. We fit these models using Bayesian linear regression,[7] where we employed an empirical Bayes approach to set model hyperparameters. Full details are provided elsewhere,[7] but this essentially consists of finding the model hyperparameters (controlling the noise- and the data variance) by maximizing the model evidence or marginal likelihood. This was achieved using conjugate gradient optimization. For fixed hyperparameters, the posterior distribution over the trend coefficients can be computed in closed form. This, in turn, enables predictions for unseen data points to be computed. We used the maximum a-posteriori estimate of the weight distribution as an indication of the importance of each trend coefficient in further analyses. To select the degree of the interpolating polynomial basis set, we fit these models across polynomials of degree 2–5 and then compared the different model orders using a Scree plot analysis.[8] This criterion strongly favoured a polynomial of degree 2 (6 TSM coefficients) for the putamen subregion and a polynomial of degree 4 (12 TSM coefficients) for the caudate-NAcc subregion. The polynomials summarized the connectivity modes well, explaining the following *mean±s.d.* of the variance: left putamen: 90.5±4.16%, right putamen: 90.2±4.64%, left caudate-NAcc: 88.6±2.54%, right caudate-NAcc: 89.4±2.15%.

### 3. Inter-subject and inter-session variability in the second-order striatal connectivity mode

The subject-specific second-order striatal connectivity modes were highly consistent across the two fMRI sessions (*mean±s.d.:* $\rho$=0.98±0.07; averaged across all four subregions), which is in line with what we have demonstrated previously for other brain regions and for the zeroth-order and first-order mode of connectivity in striatum.[4, 9] Both the variations across subjects and the reproducibility within subjects are illustrated in Figure S1 (819 of the 839 participants completed two resting-state fMRI sessions). Inter-class correlation (ICC(2,k)), which indexes measurement consistency for a putative biomarker[10, 11] also showed excellent reproducibility of the subject-specific connectivity modes for all four subregions, while still being sensitive to inter-individual differences. This was assessed through a permutation test on the session 1-to session 2 spatial correlations (N=10000) within and between subjects (Table S1).

| **Striatal subregion** | **ICC [bootstrapped 95% CI]** | **Within-subject correlation** | **Between-subject correlation** | **Within vs between permutation test** |
|---|---|---|---|---|
| left putamen | 0.960 [0.951 - 0.965] | 0.9843 | 0.9641 | $p$<0.0001 |
| left caudate-NAcc | 0.974 [0.968 - 0.978] | 0.9701 | 0.9655 | $p$=0.0941 |
| right putamen | 0.961 [0.952 - 0.967] | 0.9806 | 0.9760 | $p$=0.0251 |
| right caudate-NAcc | 0.974 [0.968 - 0.978] | 0.9812 | 0.9769 | $p$=0.0002 |

**Table S1. Interclass Correlation Coefficients (ICCs) between the two scanning sessions and the session 1-to session 2 within-subject and between-subject spatial correlations.** CI=confidence interval.

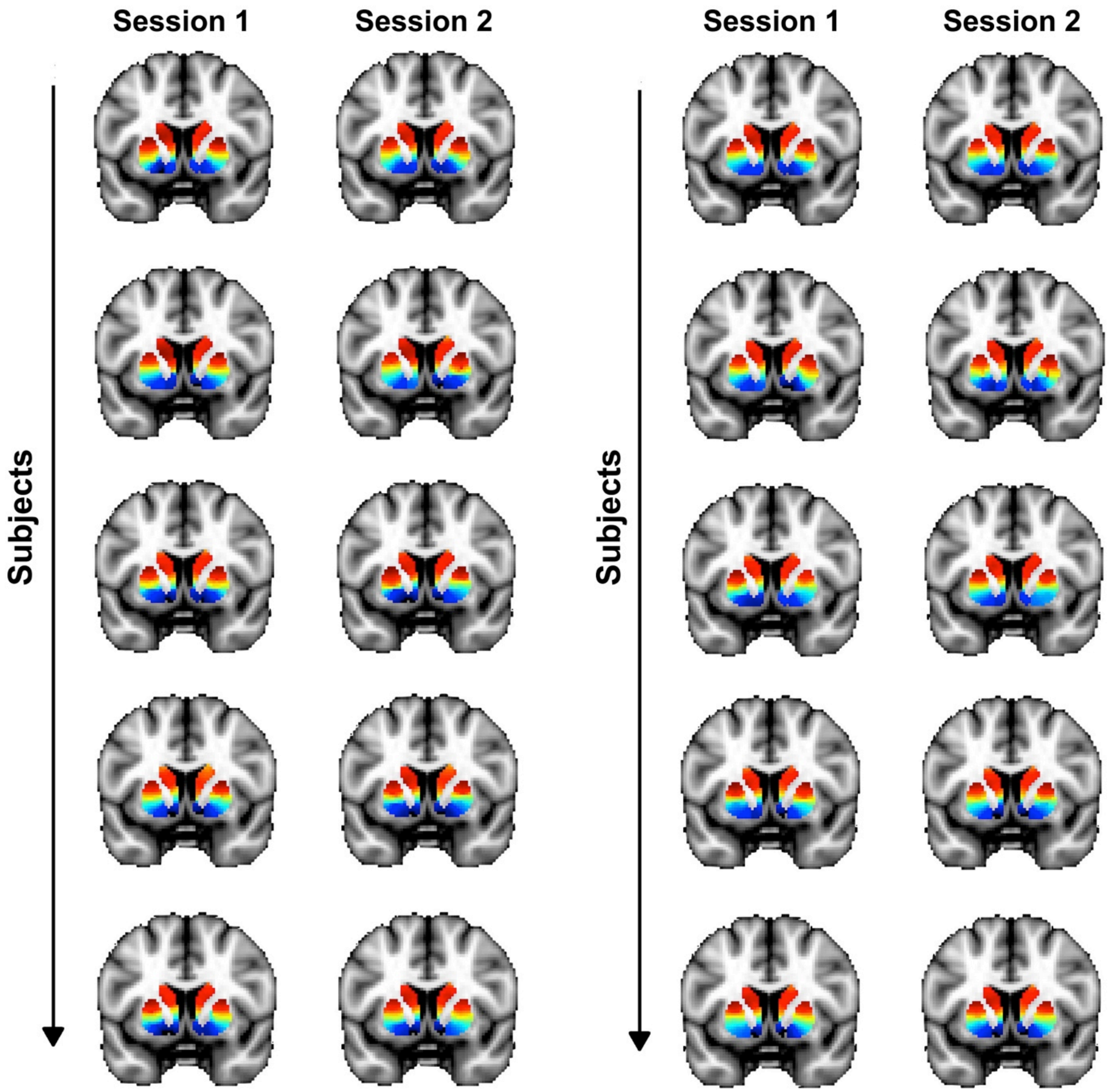

**Figure S1. Inter-subject and inter-session (within-subject) variability in the second-order mode of connectivity in striatum.** Individual-subject connectivity modes are shown for 10 randomly selected HCP subjects (from a total of 839). This figure shows variations between subjects as well as variations between sessions for the same subjects.

**4. Within-subject correspondence between second-order striatal connectivity mode and DaT SPECT scan**

In the main manuscript we demonstrated that the second-order striatal connectivity mode at the group-level (obtained by averaging this mode across all 839 HCP subjects) showed a very high spatial correlation (r=0.884) with the group-level DaT SPECT image of striatum (obtained by averaging the DAT SPECT images across all 209 PPMI controls). We also aimed to demonstrate that this mapping can be replicated at the within-subject level by investigating the within-subject spatial correspondence between this connectivity mode and the DaT SPECT scan acquired in the PPMI dataset. However, while the PPMI dataset has resting-state fMRI data available for a small subsample of its participants (14 controls with one resting-state fMRI dataset each, and 82 Parkinson's disease patients with 130 resting-state fMRI datasets combined (in case of multiple assessments per subject they were separated by at least one year), it is of a relatively low temporal and spatial resolution (*TR*=2400ms, 210 time points, 3.3mm isotropic resolution compared to the HCP data: *TR*=720ms, 2400 time points, 2.0mm isotropic resolution). While this resolution is sufficient for typical resting-state fMRI analyses, the precise delineation of the very fine-grained and overlapping connectivity modes using connectopic mapping calls for high-resolution data. The single subject connectivity modes in the PPMI dataset (as opposed to the HCP single subject modes and group-level modes) might therefore not be of sufficient quality and reliable for every subject. To address this issue we first computed the spatial correlation of each subject's individual connectivity mode with that of the group-average HCP connectivity mode as well as with the DaT SPECT scan of each subject, see Figure S2. In this analysis, the second-order striatal connectivity mode was modelled separately (and correlations were calculated separately) for the left and right putamen and caudate-NAcc subregions. This revealed highly significant positive correlations ($0.68>r<0.91$, all $p<4.0e21$) across both controls and patients, suggesting that if the connectivity mode of a subject resembles the HCP group-average connectivity mode –assumed to be an index of good quality– a high spatial similarity can be observed between the connectivity mode and the DaT SPECT scan of that subject. Next we selected those subjects with good quality connectivity modes as determined by a spatial correlation of $r>0.5$ with the group-average connectivity mode in the HCP dataset. Within this sample of 73-86 datasets from Parkinson's disease patients and 6-8 datasets from controls (dependent on the striatal subregion), we not only replicated the spatial correspondence between the connectivity mode and DaT SPECT scan at the group-level (patients: $r$=0.714; control group: $r$=0.721) but also observed *within-subject* spatial correlations of (0.44>$r$<0.63; mean=0.58, 95% CI = [0.56,0.60]) between the connectivity mode and DaT SPECT scan, see Figure S3. While we were able to replicate the spatial correlation between the second connectivity mode and the DaT SPECT scan at the *within-subject* level, this correlation ($r$=0.58) is not as high as the spatial correlations observed in the group level (i.e., $r$=0.721 and $r$=0.714 for PPMI controls and Parkinson's disease patients respectively, and $r$=0.884 between the DaT SPECT scan in PPMI controls and the connectivity mode in HCP participants). This is however not surprising given the relatively low temporal and spatial resolution of the resting-state fMRI scan of the PPMI dataset. However to our knowledge, there is currently no dataset available that includes both a high-resolution resting-state fMRI scan and a DAT-SPECT scans from the same participants.

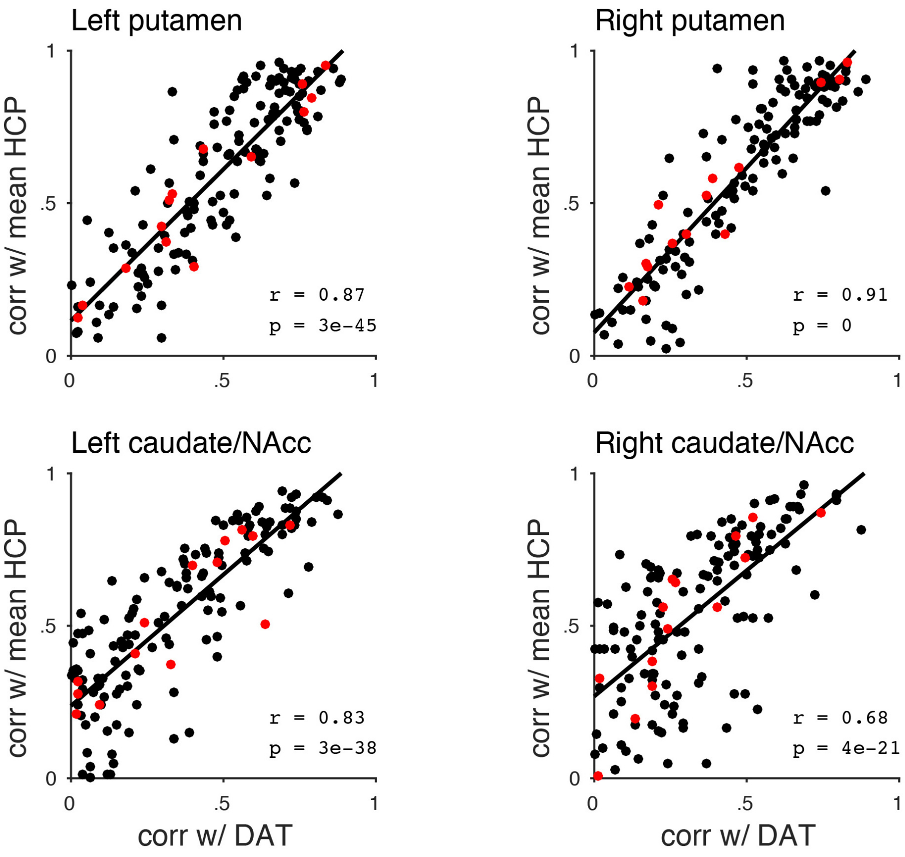

**Figure S2. Spatial correlations of the subject-specific second connectivity modes with the mean HCP connectivity mode and the DaT SPECT scan.** These plots shows that when the connectivity mode of a subject resembles the HCP group-average connectivity mode –assumed to be an index of good quality– a high spatial similarity can be observed between the connectivity mode and the DaT SPECT scan of that subject. Red dots represent control participants, black dots patients with Parkinson's disease.

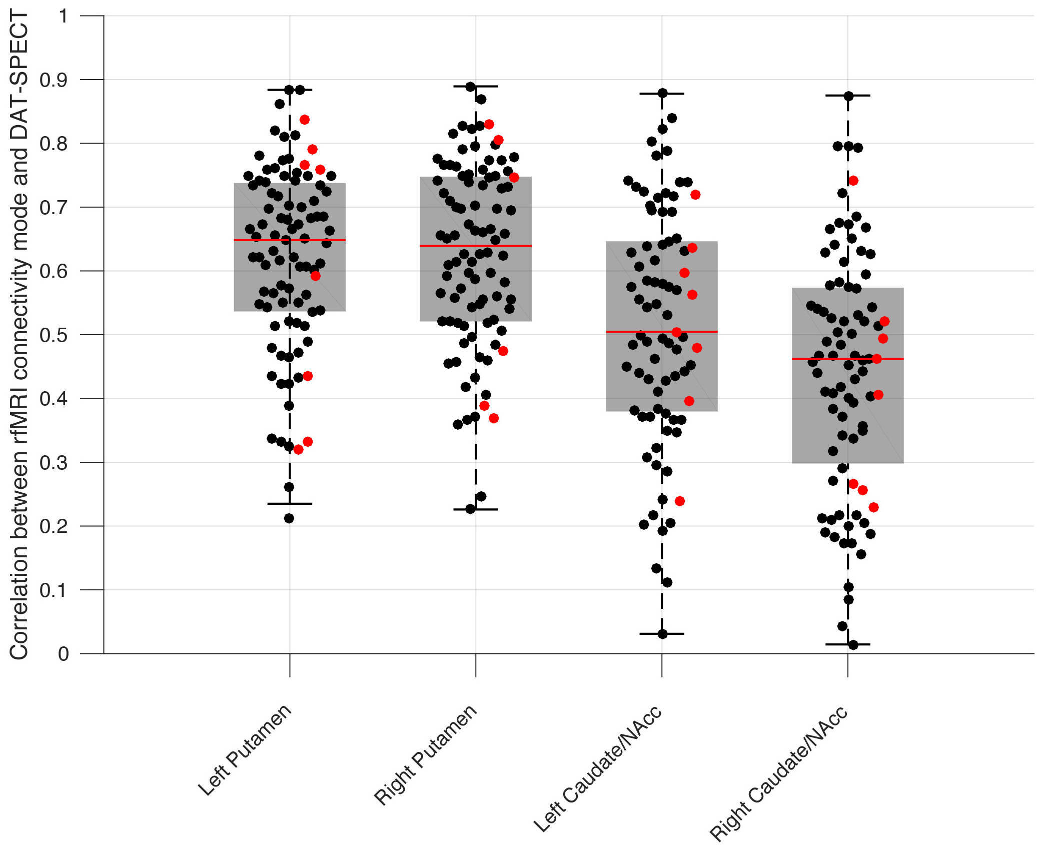

**Figure S3. Within-subject correlations between the second connectivity mode and DaT SPECT scan.** These correlations were obtained in a subsample of the PPMI dataset (6-8 datasets from controls and 73-82 datasets from Parkinson's disease patients) with connectivity modes displaying high spatial correlations ($r$>0.5) with the mean HCP connectivity mode. Red dots represent control participants, black dots represent patients with Parkinson's disease.

**5. Investigating the second-order striatal connectivity mode in Parkinson's disease**

Patients with Parkinson's Disease underwent two 10-minute resting-state fMRI sessions, i.e., a placebo session and L-DOPA session, separated by at least a day on a 3T Siemens Magnetom Prisma$^{fit}$ scanner. Resting-state fMRI scans were obtained with an interleaved high-resolution multiband sequence (*TR*=0.860s, voxel size=2.2mm isotropic, *TE*=34ms, flip angle=20°, 44 axial slices, multiband acceleration factor=4, volumes=700). Under both conditions, patients were scanned after overnight fasting in a practically defined OFF-state, i.e., more than 12h after intake of their last dose of dopaminergic medication. During one session patients were scanned after administration of L-DOPA, i.e., they received a standardized dose of 200/50 mg dispersible levodopa/benserazide. During the other session patients received placebo (cellulose powder). The cellulose powder and L-DOPA/benserazide were dissolved in water and therefore undistinguishable for the participants. Patients also received 10 mg domperidone to improve gastro-intestinal absorption of levodopa and reduce side effects. The order of sessions was counterbalanced and the resting-state fMRI scan started on average 48 min (range: 25-70 min) after taking L-DOPA or placebo. Symptom severity was assessed during both sessions with part III (assessment of motor function by a clinician) of the Movement Disorders Society Unified Parkinson Disease Rating Scale (UPDRS)[12] and an electromyogram (EMG) of the hand was recorded to monitor tremor-related activity. In light of ethical considerations, control participants did not receive L-DOPA and placebo, they just underwent two typical resting-state fMRI sessions during which the UPDRS was not administered.

Preprocessing of the resting-state fMRI data included removal of the first five volumes to allow for signal equilibration, primary head motion correction via realignment to the middle volume MCFLIRT,[13] grand mean scaling, and spatial smoothing with a 6mm FWHM Gaussian kernel. The pre-processing pipeline was furthermore designed to rigorously correct for potential tremor-induced head-motion related artefacts. To this end, we used ICA-AROMA,[14] an advanced ICA-based motion correction procedure to identify and remove secondary head motion-related artefacts with high accuracy while preserving signal of interest.[14,15] Next, any remaining motion artefacts were removed from the data by regressing out the EMG parameters in addition to the white matter and CSF signal.[16] Finally, the data were temporally filtered with a high-pass filter of 0.01Hz before being resampled to 2mm MNI152 space.

We applied connectopic mapping to the pre-processed resting-state fMRI data of each session from every participant and selected the second-order connectivity mode for further analyses, using the same procedure as in the HCP dataset. The subject-specific second-order striatal connectivity modes for control participants were again consistent across the two fMRI sessions *mean±s.d.* $\rho$=0.85±0.11 (individual subregions: left putamen: $\rho$=0.78±0.10, right putamen: $\rho$=0.82±0.12, left caudate-NAcc: $\rho$=0.87±0.14, and right caudate-NAcc: $\rho$=0.92±0.08). The polynomials also summarized the connectivity modes well, explaining mean±s.d. 78.6±11.8% of the variance across the striatum in controls (individual subregions: left putamen: 67.9±16.2%, right putamen: 65.3±21.2%, left caudate-NAcc: 90.5±4.28%, right caudate-NAcc: 90.8±5.63%), and explaining *mean±s.d.* 78.0±10.5% of the variance across striatum in Parkinson's disease patients under placebo (individual subregions: left putamen: 63.6±19.6%, right putamen: 69.5±13.0%, left caudate-NAcc: 88.5±4.69%, right caudate-NAcc: 90.4±4.58%). While these numbers are lower than observed for the connectivity modes obtained from the HCP dataset – which is not surprising given the exceptionally high quality of the HCP dataset– the reproducibility and explained variance of the TSM coefficients is still substantial.

### 6. Post-hoc correlations with tobacco and alcohol use for all significant TSM coefficients

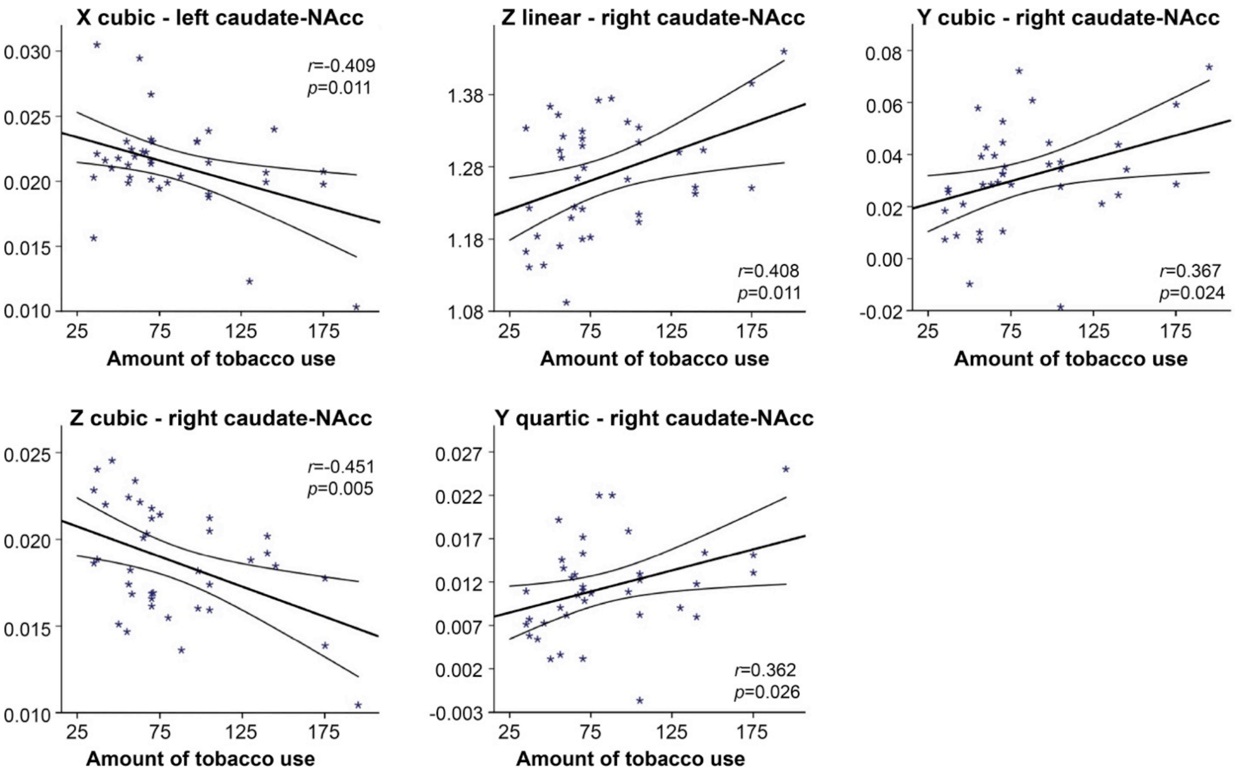

**Figure S4. The second-order mode of connectivity in striatum is associated with the amount of tobacco use.** A strong association was observed between the TSM coefficients modelling the connectivity mode in the caudate-NAcc region and the total amount of tobacco use over the past week (GLM omnibus test: $X^2$=49.55, $p$=0.002). To visualize this relationship, Pearson correlations between the individual TSM coefficients and the amount of use were computed and the correlations reaching significance ($p$<0.05) are shown in this figure.

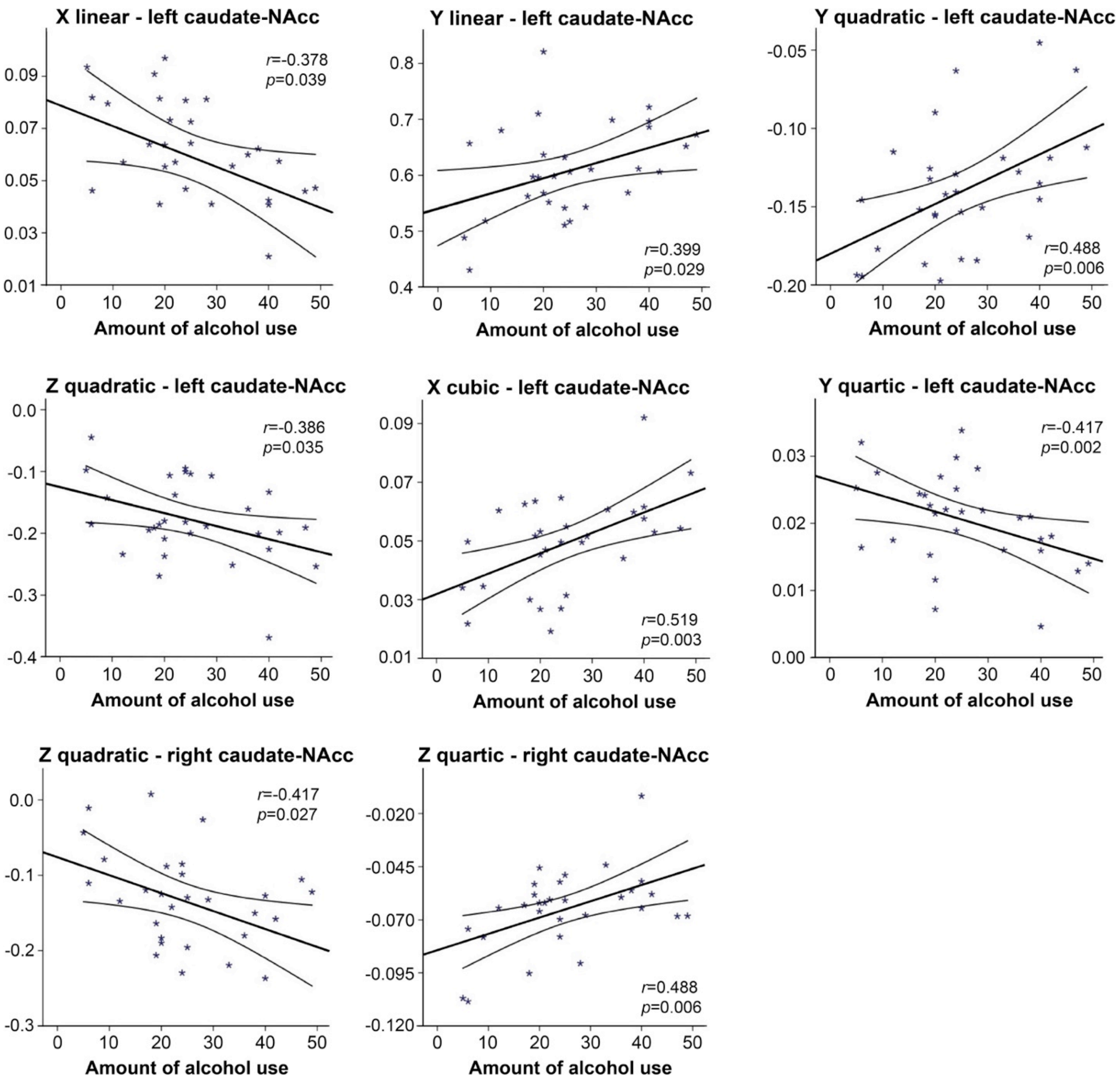

**Figure S5. The second-order mode of connectivity in striatum is associated with the amount of alcohol use.** A strong association was observed between the TSM coefficients modelling the connectivity mode in the caudate-NAcc region and the total number of alcoholic drinks over the past week (GLM omnibus test: $X^2$=64.45, $p$<0.001). To visualize this relationship, Pearson correlations between the individual TSM coefficients and the amount of use were computed and the correlations reaching significance ($p$<0.05) are shown in this figure.

### 7. Post-hoc analyses of age and sex

For all the analyses described in the main manuscript (effects of diagnosis and L-DOPA in the Parkinson's disease dataset and associations with smoking and drinking in the HCP dataset), we conducted post-hoc sensitivity analyses to rule out that the group differences and behavioural associations revealed by our analyses were dependent on age and sex. To this end, we conducted two types of analyses. First we repeated our main analyses by including covariates for age and sex in our statistical models in addition to the TSM coefficients, to verify that effects remained (close to) significant when including these demographic variables. Next, we only included age and sex in our statistical models (without the TSM coefficients) to verify that effects were not explained by age and/or sex only. The outcomes of these analyses ($X^2$ and $p$-value) are listed in Table S2 and demonstrate that none of the significant effects observed in our main analyses were dependent on age or sex. However, adding age and sex (age in particular) did increase the significance of findings substantially for the analyses investigating the L-DOPA induced changes. This might be explained by the fact that patients who are older often have more severe Parkinson's disease and do not benefit as much anymore from L-DOPA treatment.

| | **Original analysis** | | **Original analysis + Age & sex** | | **Age & sex only** | |
|---|---|---|---|---|---|---|
| | $X^2$ | *p-value* | $X^2$ | *p-value* | $X^2$ | *p-value* |
| *PUTAMEN* | | | | | | |
| **Patients vs controls** right tremor-dominant Parkinson's disease | 27.17 | *0.007* | 27.21 | *0.018* | 0.48 | *0.786* |
| **UPDRS symptom severity** right tremor-dominant Parkinson's disease | 22.28 | *0.035* | 23.46 | *0.053* | 2.38 | *0.305* |
| **L-DOPA-placebo difference** left tremor-dominant Parkinson's disease | 34.07 | *0.001* | 46.14 | *<0.001* | 2.42 | *0.299* |
| **L-DOPA-placebo difference** right tremor-dominant Parkinson's disease | 25.48 | *0.012* | 37.53 | *0.001* | 7.18 | *0.028* |
| *CAUDATE-NACC* | | | | | | |
| **Tobacco use** HCP dataset | 49.55 | *0.002* | 53.56 | *0.001* | 1.04 | *0.594* |
| **Alcohol use** HCP dataset | 64.45 | *<0.001* | 174.87 | *<0.001* | 9.26 | *0.010* |

**Table S2. Post-hoc analyses of age and sex.**

### 8. Post-hoc analyses using different usage thresholds for tobacco and alcohol use

We also investigated whether the associations of the second-order mode of connectivity in striatum with the amount of tobacco use and alcohol use persisted under different usage thresholds. For both tobacco and alcohol use we chose a daily usage threshold lower (≥2x tobacco/≥1x alcoholic drink) and a daily usage threshold higher (≥8x tobacco/≥3x alcoholic drink) than the one used in the main analysis (≥5x tobacco/≥3x light alcoholic and/or ≥1x hard liquor drinks a day). Please note that the aim of these analyses is not necessarily to show that effects remain significant as under different usage thresholds the sample size and statistical power will change, but rather that the explained variance remains high. Nevertheless, apart from the low usage threshold for alcohol use, all effects also remained significant, as can be observed in Tables S3 and S4, indicating that the associations with tobacco and alcohol use were not specific to the chosen usage threshold. However, a pattern that is visible is that associations become stronger when only including the highest users in this population-based sample in the analysis.

| | **Original analysis:<br>≥5x tobacco use a day**<br>*N*=38 | | **≥2x tobacco use a day**<br>*N*=62 | | **≥8x tobacco use a day**<br>*N*=30 | |
|---|---|---|---|---|---|---|
| | $X^2$ | *p-value* | $X^2$ | *p-value* | $X^2$ | *p-value* |
| **Tobacco use**<br>HCP dataset<br>*caudate-NAcc* | 49.55 | *0.002* | 37.96 | *0.035* | 70.54 | *<0.001* |

**Table S3. Post-hoc analyses using different thresholds for tobacco use.**

| | **Original analysis:<br>≥3x light alcoholic and/or<br>≥1x hard liquor drinks a day**<br>*N*=30 | | **≥1x alcoholic drinks a day (light and/or hard liquor)**<br>*N*=103 | | **≥3x alcoholic drinks a day (light and/or hard liquor) ***<br>*N*=26 | |
|---|---|---|---|---|---|---|
| | $X^2$ | *p-value* | $X^2$ | *p-value* | $X^2$ | *p-value* |
| **Alcohol use**<br>HCP dataset<br>*caudate-NAcc* | 64.45 | *<0.001* | 29.94 | *0.187* | 196.57 | *<0.001* |

**Table S4. Post-hoc analyses using different thresholds for alcohol use.**

### 9. References

**1.** Van Essen DC, Smith SM, Barch DM, et al. The WU-Minn human connectome project: an overview. *Neuroimage.* 2013;80:62-79.

**2.** Glasser MF, Sotiropoulos SN, Wilson JA, et al. The minimal preprocessing pipelines for the Human Connectome Project. *Neuroimage.* 2013;80:105-124.

**3.** Salimi-Khorshidi G, Douaud G, Beckmann CF, Glasser MF, Griffanti L, Smith SM. Automatic denoising of functional MRI data: combining independent component analysis and hierarchical fusion of classifiers. *Neuroimage.* 2014;90:449-468.

**4.** Haak KV, Marquand AF, Beckmann CF. Connectopic mapping with resting-state fMRI. *NeuroImage.* 2018/04/15/ 2018;170:83-94.

**5.** Belkin M, Niyogi P. Laplacian eigenmaps and spectral techniques for embedding and clustering. Paper presented at: Advances in neural information processing systems, 2002.

**6.** Gelfand AE, Diggle P, Guttorp P, Fuentes M. *Handbook of spatial statistics*: CRC press; 2010.

**7.** Bishop CM. Graphical models. *Pattern recognition and machine learning.* 2006;4:359-422.

**8.** Cattell RB. The scree test for the number of factors. *Multivariate behavioral research.* 1966;1(2):245-276.

**9.** Marquand AF, Haak KV, Beckmann CF. Functional corticostriatal connection topographies predict goal-directed behaviour in humans. *Nature human behaviour.* 2017;1:s41562-41017-40146.

**10.** Shrout PE, Fleiss JL. Intraclass correlations: uses in assessing rater reliability. *Psychological bulletin.* 1979;86(2):420.

**11.** Koo TK, Li MY. A guideline of selecting and reporting intraclass correlation coefficients for reliability research. *Journal of chiropractic medicine.* 2016;15(2):155-163.

**12.** Goetz CG, Tilley BC, Shaftman SR, et al. Movement Disorder Society-sponsored revision of the Unified Parkinson's Disease Rating Scale (MDS-UPDRS): scale presentation and clinimetric testing results. *Movement disorders: official journal of the Movement Disorder Society.* 2008;23(15):2129-2170.

**13.** Jenkinson M, Bannister P, Brady M, Smith S. Improved optimization for the robust and accurate linear registration and motion correction of brain images. *Neuroimage.* 2002;17(2):825-841.

**14.** Pruim RH, Mennes M, Buitelaar JK, Beckmann CF. Evaluation of ICA-AROMA and alternative strategies for motion artifact removal in resting state fMRI. *Neuroimage.* 2015;112:278-287.

**15.** Parkes L, Fulcher B, Yu M, Fornitod A. An evaluation of the efficacy, reliability, and sensitivity of motion correction strategies for resting-state functional MRI. *NeuroImage.* 2017.

**16.** Helmich RC, Bloem BR, Toni I. Motor imagery evokes increased somatosensory activity in Parkinson's disease patients with tremor. *Human Brain Mapping.* 2012;33(8):1763-1779.

### 10. Subject IDs from all HCP and PPMI subjects included in our analyses

**Table S5:** Subject IDs from the 839 HCP subjects used in our connectoppic mapping analysis.

| | | | | | | | |
|---|---|---|---|---|---|---|---|
| 100206 | 129129 | 155635 | 181636 | 212823 | 385450 | 580044 | 784565 |
| 100610 | 129331 | 155938 | 182032 | 213017 | 386250 | 580347 | 788674 |
| 101006 | 129533 | 156031 | 182436 | 213421 | 387959 | 580650 | 789373 |
| 101107 | 129634 | 156435 | 183034 | 213522 | 389357 | 580751 | 792766 |
| 101309 | 129937 | 156536 | 183337 | 214524 | 391748 | 581450 | 792867 |
| 101410 | 130114 | 157437 | 183741 | 214625 | 392447 | 583858 | 793465 |
| 101915 | 130316 | 157942 | 185341 | 214726 | 392750 | 585256 | 800941 |
| 102008 | 130417 | 158136 | 185442 | 217126 | 393247 | 587664 | 802844 |
| 102109 | 130619 | 158338 | 185846 | 219231 | 393550 | 588565 | 803240 |
| 102311 | 130720 | 158843 | 185947 | 220721 | 394956 | 589567 | 804646 |
| 102513 | 130821 | 159138 | 186040 | 221218 | 395251 | 590047 | 809252 |
| 102614 | 131217 | 159340 | 186141 | 223929 | 395756 | 592455 | 810439 |
| 102715 | 131419 | 159441 | 186545 | 227432 | 395958 | 594156 | 810843 |
| 103010 | 131722 | 159744 | 186848 | 227533 | 397154 | 597869 | 812746 |
| 103111 | 131823 | 159845 | 187143 | 228434 | 397861 | 599065 | 814548 |
| 103212 | 132017 | 159946 | 187345 | 231928 | 406432 | 599469 | 814649 |
| 104012 | 132118 | 160729 | 187547 | 233326 | 406836 | 599671 | 815247 |
| 104416 | 133019 | 160830 | 187850 | 236130 | 412528 | 601127 | 816653 |
| 104820 | 134021 | 160931 | 188145 | 237334 | 413934 | 604537 | 818455 |
| 105014 | 134223 | 161630 | 188347 | 238033 | 419239 | 609143 | 818859 |
| 105620 | 134425 | 161832 | 188448 | 239136 | 421226 | 611938 | 820745 |
| 105923 | 134627 | 162026 | 188549 | 248339 | 422632 | 613235 | 822244 |
| 106016 | 134829 | 162228 | 188751 | 250932 | 424939 | 613538 | 825048 |
| 106521 | 135124 | 162733 | 189349 | 255740 | 432332 | 615441 | 825553 |
| 106824 | 135225 | 162935 | 189450 | 256540 | 436239 | 615744 | 825654 |
| 107018 | 135528 | 163129 | 191033 | 257542 | 436845 | 616645 | 826454 |
| 107220 | 135629 | 163331 | 191235 | 257845 | 441939 | 617748 | 827052 |
| 107321 | 135730 | 163836 | 191336 | 257946 | 445543 | 618952 | 828862 |
| 107422 | 136126 | 164030 | 191841 | 263436 | 449753 | 620434 | 832651 |
| 107725 | 136227 | 164131 | 191942 | 268749 | 453441 | 622236 | 833148 |
| 108020 | 136631 | 164636 | 192035 | 268850 | 453542 | 623137 | 833249 |
| 108121 | 136732 | 164939 | 192136 | 270332 | 454140 | 623844 | 835657 |
| 108222 | 137027 | 165032 | 192237 | 274542 | 456346 | 626648 | 837560 |
| 108323 | 137229 | 165234 | 192641 | 275645 | 459453 | 627852 | 837964 |
| 108525 | 137431 | 165436 | 192843 | 280739 | 461743 | 633847 | 841349 |
| 108828 | 137532 | 165638 | 193441 | 280941 | 463040 | 634748 | 843151 |
| 109123 | 137633 | 165941 | 193845 | 281135 | 467351 | 635245 | 844961 |
| 109325 | 137936 | 166438 | 194443 | 283543 | 468050 | 644044 | 845458 |
| 109830 | 138130 | 166640 | 194645 | 285345 | 473952 | 645450 | 849264 |
| 110007 | 138332 | 167036 | 194746 | 285446 | 475855 | 647858 | 849971 |
| 111211 | 138837 | 167238 | 194847 | 286347 | 479762 | 654350 | 852455 |
| 111413 | 139233 | 167440 | 195041 | 286650 | 480141 | 654552 | 856463 |
| 112112 | 139435 | 168240 | 195445 | 287248 | 481042 | 656253 | 856968 |
| 112314 | 139839 | 168341 | 195950 | 289555 | 481951 | 657659 | 867468 |
| 112516 | 140117 | 168745 | 196346 | 290136 | 486759 | 660951 | 869472 |
| 112920 | 140319 | 168947 | 196851 | 295146 | 492754 | 662551 | 870861 |
| 113316 | 140824 | 169040 | 196952 | 297655 | 495255 | 663755 | 871762 |

| | | | | | | | |
|---|---|---|---|---|---|---|---|
| 113922 | 140925 | 169444 | 197348 | 298455 | 497865 | 664757 | 872562 |
| 114116 | 141119 | 169545 | 197651 | 299154 | 500222 | 667056 | 873968 |
| 114217 | 141422 | 169747 | 198047 | 299760 | 506234 | 668361 | 877269 |
| 114318 | 141826 | 169949 | 198249 | 300618 | 510225 | 671855 | 878776 |
| 114419 | 142424 | 170631 | 198350 | 300719 | 510326 | 673455 | 878877 |
| 114621 | 143224 | 170934 | 198653 | 303119 | 512835 | 675661 | 880157 |
| 114823 | 143426 | 171128 | 198855 | 303624 | 513130 | 677766 | 882161 |
| 115017 | 144125 | 171330 | 199352 | 304727 | 513736 | 679568 | 884064 |
| 115219 | 144731 | 171431 | 199453 | 305830 | 516742 | 679770 | 886674 |
| 115724 | 144832 | 171532 | 200008 | 308129 | 517239 | 680250 | 888678 |
| 115825 | 144933 | 171633 | 200109 | 308331 | 518746 | 680452 | 891667 |
| 116221 | 145127 | 171734 | 200311 | 309636 | 519647 | 683256 | 894067 |
| 116423 | 145531 | 172029 | 200513 | 310621 | 519950 | 686969 | 894774 |
| 116524 | 145632 | 172130 | 200917 | 311320 | 520228 | 687163 | 898176 |
| 116726 | 145834 | 172433 | 201414 | 314225 | 521331 | 689470 | 901038 |
| 117021 | 146129 | 172534 | 201717 | 316633 | 522434 | 690152 | 901442 |
| 117728 | 146331 | 172635 | 201818 | 316835 | 523032 | 692964 | 902242 |
| 117930 | 146432 | 172938 | 202113 | 317332 | 524135 | 693764 | 905147 |
| 118023 | 146533 | 173132 | 202719 | 318637 | 525541 | 694362 | 907656 |
| 118124 | 146634 | 173334 | 203418 | 320826 | 529549 | 695768 | 908860 |
| 118225 | 146735 | 173435 | 203923 | 321323 | 529953 | 698168 | 910241 |
| 118528 | 146937 | 173536 | 204016 | 322224 | 531536 | 700634 | 910443 |
| 118831 | 147030 | 173637 | 204319 | 325129 | 536647 | 701535 | 911849 |
| 119025 | 147636 | 173738 | 204420 | 329844 | 540436 | 706040 | 912447 |
| 119126 | 147737 | 173839 | 204521 | 330324 | 541640 | 707749 | 917558 |
| 119732 | 148133 | 173940 | 204622 | 333330 | 545345 | 715041 | 919966 |
| 120414 | 148335 | 174841 | 205220 | 334635 | 547046 | 715950 | 922854 |
| 120515 | 148436 | 175136 | 206222 | 339847 | 548250 | 720337 | 923755 |
| 120717 | 148941 | 175237 | 206323 | 341834 | 549757 | 724446 | 926862 |
| 121315 | 149236 | 175338 | 206525 | 342129 | 550439 | 725751 | 927359 |
| 121416 | 149741 | 175540 | 206727 | 346137 | 552241 | 727553 | 929464 |
| 121618 | 149842 | 175742 | 206828 | 346945 | 553344 | 727654 | 930449 |
| 121921 | 150625 | 176037 | 206929 | 348545 | 555348 | 728454 | 933253 |
| 122317 | 150726 | 176441 | 207123 | 349244 | 555651 | 729254 | 942658 |
| 122418 | 150928 | 176744 | 207426 | 350330 | 555954 | 731140 | 947668 |
| 122620 | 151021 | 176845 | 208024 | 352132 | 557857 | 734247 | 952863 |
| 122822 | 151324 | 177140 | 208125 | 352738 | 558657 | 735148 | 953764 |
| 123420 | 151425 | 177241 | 208327 | 353740 | 558960 | 737960 | 955465 |
| 123521 | 151728 | 177342 | 208428 | 355239 | 559457 | 742549 | 957974 |
| 123723 | 151829 | 177645 | 208630 | 356948 | 561444 | 744553 | 958976 |
| 123824 | 151930 | 178142 | 209127 | 358144 | 561949 | 748662 | 962058 |
| 123925 | 152225 | 178243 | 209228 | 360030 | 562345 | 749058 | 965771 |
| 124220 | 152427 | 178647 | 209329 | 361234 | 562446 | 751550 | 966975 |
| 124624 | 152831 | 178748 | 209531 | 361941 | 565452 | 753150 | 970764 |
| 124826 | 153025 | 178849 | 209834 | 362034 | 566454 | 757764 | 971160 |
| 125222 | 153126 | 178950 | 210011 | 365343 | 567052 | 759869 | 972566 |
| 125424 | 153227 | 179245 | 210112 | 366042 | 567961 | 760551 | 973770 |
| 126426 | 153631 | 179346 | 210415 | 368551 | 568963 | 763557 | 978578 |
| 126628 | 153732 | 179952 | 211114 | 368753 | 569965 | 765864 | 979984 |
| 127226 | 153833 | 180129 | 211215 | 376247 | 571144 | 766563 | 983773 |
| 127327 | 153934 | 180230 | 211316 | 377451 | 571548 | 769064 | 987074 |
| 127630 | 154229 | 180432 | 211619 | 378756 | 572045 | 770352 | 989987 |
| 127731 | 154330 | 180533 | 211821 | 378857 | 573249 | 771354 | 990366 |
| 127832 | 154532 | 180735 | 211922 | 379657 | 573451 | 773257 | 991267 |

| | | | | | | | |
|---|---|---|---|---|---|---|---|
| 128026 | 154734 | 180836 | 212015 | 380036 | 576255 | 774663 | 992673 |
| 128127 | 154835 | 180937 | 212116 | 381038 | 578057 | 779370 | 993675 |
| 128329 | 154936 | 181131 | 212217 | 381543 | 578158 | 782561 | 996782 |
| 128935 | 155231 | 181232 | 212419 | 382242 | 579867 | 783462 | 788674 |

**Table S6:** The 209 PPMI controls with DaT SPECT data used in our analysis.

| PPMI Subject ID | Image ID DaT SPECT | PPMI Subject ID | Image ID DaT SPECT | PPMI Subject ID | Image ID DaT SPECT |
|---|---|---|---|---|---|
| 3000 | 323662 | 3350 | 339901 | 3637 | 388521 |
| 3004 | 341194 | 3351 | 339902 | 3639 | 388523 |
| 3008 | 341195 | 3353 | 339904 | 3651 | 339008 |
| 3009 | 341196 | 3355 | 341236 | 3651 | 355956 |
| 3011 | 341198 | 3357 | 339907 | 3656 | 339014 |
| 3013 | 341200 | 3358 | 339908 | 3658 | 339016 |
| 3016 | 341202 | 3361 | 339911 | 3662 | 355221 |
| 3029 | 388468 | 3362 | 339912 | 3668 | 388528 |
| 3053 | 341207 | 3363 | 338780 | 3750 | 388535 |
| 3055 | 341209 | 3368 | 339917 | 3754 | 360616 |
| 3057 | 341211 | 3369 | 339918 | 3756 | 360617 |
| 3064 | 341217 | 3370 | 339919 | 3759 | 363950 |
| 3069 | 341221 | 3389 | 388504 | 3765 | 363951 |
| 3070 | 341222 | 3390 | 388505 | 3767 | 388536 |
| 3071 | 341223 | 3401 | 340345 | 3768 | 363952 |
| 3072 | 341224 | 3404 | 340346 | 3769 | 360618 |
| 3073 | 341225 | 3405 | 340347 | 3779 | 453700 |
| 3074 | 341226 | 3410 | 340351 | 3794 | 388545 |
| 3075 | 341227 | 3411 | 340352 | 3796 | 388147 |
| 3085 | 388470 | 3414 | 340354 | 3803 | 355230 |
| 3087 | 388472 | 3424 | 340363 | 3804 | 354344 |
| 3100 | 341230 | 3438 | 340388 | 3805 | 354345 |
| 3103 | 341233 | 3450 | 340398 | 3806 | 354346 |
| 3104 | 339536 | 3452 | 339923 | 3807 | 355231 |
| 3106 | 340418 | 3453 | 339924 | 3811 | 360620 |
| 3109 | 340423 | 3457 | 339928 | 3812 | 355232 |
| 3112 | 340426 | 3458 | 339929 | 3813 | 355233 |
| 3114 | 340430 | 3460 | 341243 | 3816 | 363953 |
| 3115 | 340431 | 3464 | 341245 | 3817 | 388148 |
| 3151 | 341018 | 3466 | 339932 | 3850 | 337832 |
| 3156 | 341021 | 3468 | 339934 | 3851 | 337833 |
| 3157 | 341022 | 3478 | 360613 | 3852 | 337834 |
| 3160 | 341023 | 3479 | 363945 | 3853 | 337835 |
| 3161 | 341024 | 3480 | 388509 | 3854 | 337836 |
| 3165 | 341027 | 3481 | 388510 | 3855 | 337445 |
| 3169 | 341031 | 3503 | 340400 | 3857 | 337837 |
| 3171 | 341033 | 3515 | 340408 | 3859 | 337839 |
| 3172 | 341034 | 3517 | 341248 | 3907 | 388556 |
| 3188 | 388483 | 3518 | 339537 | 3908 | 363957 |
| 3191 | 388486 | 3521 | 339539 | 3917 | 388563 |
| 3200 | 341036 | 3523 | 339541 | 3950 | 341083 |
| 3201 | 341037 | 3524 | 339542 | 3952 | 341085 |
| 3202 | 341038 | 3525 | 339543 | 3955 | 388565 |
| 3204 | 341040 | 3526 | 339544 | 3959 | 355241 |
| 3206 | 341042 | 3527 | 339545 | 3965 | 388573 |
| 3208 | 341044 | 3541 | 355215 | 3966 | 388574 |

| | | | | | |
|---|---|---|---|---|---|
| 3213 | 341049 | 3543 | 363946 | 3967 | 388576 |
| 3215 | 341051 | 3544 | 388514 | 3968 | 388577 |
| 3216 | 341052 | 3551 | 339550 | 3969 | 388578 |
| 3217 | 341053 | 3554 | 339552 | 4004 | 339032 |
| 3219 | 341055 | 3554 | 358138 | 4007 | 339035 |
| 3221 | 341057 | 3555 | 339553 | 4008 | 339036 |
| 3222 | 341058 | 3563 | 339559 | 4009 | 339037 |
| 3235 | 388488 | 3565 | 339561 | 4010 | 339038 |
| 3237 | 388490 | 3569 | 339564 | 4014 | 389268 |
| 3257 | 341067 | 3570 | 339565 | 4018 | 339045 |
| 3260 | 341068 | 3571 | 389245 | 4032 | 388583 |
| 3264 | 341070 | 3572 | 338781 | 4063 | 355246 |
| 3270 | 341074 | 3576 | 338785 | 4067 | 388593 |
| 3271 | 341075 | 3600 | 338788 | 4079 | 388596 |
| 3274 | 341077 | 3611 | 338797 | 4090 | 343886 |
| 3276 | 341079 | 3613 | 338799 | 4095 | 354353 |
| 3277 | 388491 | 3614 | 338800 | 4100 | 360623 |
| 3286 | 388494 | 3615 | 338801 | 4104 | 363963 |
| 3300 | 339889 | 3619 | 339001 | 4105 | 388600 |
| 3301 | 339890 | 3620 | 339002 | 4116 | 388613 |
| 3310 | 339896 | 3624 | 341251 | 4118 | 388615 |
| 3316 | 342187 | 3627 | 342204 | 4139 | 388627 |
| 3318 | 342189 | 3635 | 388519 | 4140 | 388628 |
| 3320 | 342191 | 3636 | 388520 | | |

**Table S7:** Patients with Parkinson's disease and controls with resting-state fMRI data & DaT SPECT data from the PPMI dataset used in our analysis.

| PPMI Subject ID | Image ID DaT SPECT | Image ID MRI | Diagnosis |
|---|---|---|---|
| 3310 | 339896 | 369414 | Control |
| 3318 | 342189 | 374882 | Control |
| 3350 | 339901 | 515208 | Control |
| 3351 | 339902 | 508245 | Control |
| 3353 | 339904 | 515216 | Control |
| 3361 | 339911 | 581042 | Control |
| 3369 | 339918 | 544617 | Control |
| 3389 | 388504 | 367349 | Control |
| 3551 | 339550 | 548987 | Control |
| 3563 | 339559 | 548989 | Control |
| 3565 | 339561 | 560369 | Control |
| 3769 | 360618 | 362609 | Control |
| 4018 | 339045 | 365285 | Control |
| 4032 | 388583 | 367390 | Control |
| 3107 | 419849 | 378215 | PD |
| 3108 | 419850 | 378223 | PD |
| 3116 | 418649 | 366137 | PD |
| 3116 | 419854 | 417052 | PD |
| 3118 | 418470 | 362555 | PD |
| 3118 | 446107 | 430138 | PD |
| 3119 | 418650 | 382277 | PD |
| 3119 | 446108 | 430147 | PD |
| 3120 | 418651 | 374854 | PD |
| 3122 | 419241 | 382284 | PD |
| 3123 | 418652 | 382289 | PD |
| 3123 | 449008 | 440114 | PD |
| 3124 | 418653 | 387304 | PD |
| 3124 | 449009 | 440118 | PD |
| 3125 | 418654 | 387314 | PD |
| 3125 | 449010 | 440128 | PD |
| 3126 | 418655 | 397752 | PD |
| 3126 | 449011 | 440131 | PD |
| 3128 | 419553 | 395434 | PD |
| 3128 | 504427 | 466848 | PD |
| 3130 | 360608 | 355962 | PD |
| 3130 | 419554 | 417000 | PD |
| 3132 | 436066 | 423718 | PD |
| 3132 | 504428 | 498892 | PD |
| 3134 | 388480 | 369013 | PD |
| 3134 | 436067 | 436351 | PD |
| 3327 | 389212 | 362478 | PD |
| 3327 | 486550 | 412180 | PD |
| 3332 | 388500 | 378540 | PD |
| 3352 | 418905 | 372319 | PD |
| 3354 | 418906 | 372327 | PD |
| 3359 | 419866 | 397593 | PD |
| 3360 | 419867 | 393662 | PD |
| 3364 | 419868 | 393672 | PD |
| 3365 | 419659 | 397597 | PD |

| | | | |
|---|---|---|---|
| 3366 | 419869 | 397624 | PD |
| 3367 | 419870 | 393674 | PD |
| 3371 | 418673 | 365166 | PD |
| 3372 | 436070 | 369487 | PD |
| 3372 | 446121 | 420330 | PD |
| 3373 | 418674 | 387316 | PD |
| 3373 | 449019 | 440174 | PD |
| 3374 | 418675 | 393614 | PD |
| 3374 | 446122 | 430165 | PD |
| 3377 | 418677 | 393628 | PD |
| 3377 | 449020 | 440186 | PD |
| 3378 | 418678 | 387324 | PD |
| 3380 | 418679 | 393636 | PD |
| 3380 | 468270 | 449575 | PD |
| 3383 | 355208 | 351070 | PD |
| 3383 | 419560 | 415707 | PD |
| 3385 | 360612 | 353398 | PD |
| 3385 | 436861 | 415713 | PD |
| 3386 | 388502 | 369048 | PD |
| 3387 | 389214 | 357590 | PD |
| 3387 | 436071 | 417033 | PD |
| 3392 | 388507 | 372995 | PD |
| 3392 | 442969 | 436390 | PD |
| 3552 | 418922 | 378354 | PD |
| 3556 | 418923 | 372348 | PD |
| 3556 | 504848 | 482323 | PD |
| 3557 | 504849 | 482329 | PD |
| 3559 | 418926 | 372359 | PD |
| 3559 | 504850 | 491605 | PD |
| 3574 | 419676 | 414623 | PD |
| 3575 | 419677 | 581115 | PD |
| 3575 | 418690 | 365225 | PD |
| 3585 | 449026 | 440198 | PD |
| 3586 | 468275 | 449581 | PD |
| 3587 | 468276 | 449584 | PD |
| 3591 | 388516 | 373018 | PD |
| 3591 | 504435 | 491626 | PD |
| 3592 | 388517 | 373035 | PD |
| 3592 | 442973 | 436404 | PD |
| 3593 | 388518 | 369096 | PD |
| 3593 | 436073 | 430199 | PD |
| 3593 | 504436 | 507400 | PD |
| 3758 | 418698 | 374893 | PD |
| 3758 | 419880 | 402067 | PD |
| 3760 | 418499 | 362591 | PD |
| 3787 | 419576 | 412194 | PD |
| 3800 | 389258 | 393684 | PD |
| 3808 | 419885 | 402071 | PD |
| 3815 | 419886 | 581145 | PD |
| 3818 | 446139 | 440242 | PD |
| 3819 | 419270 | 395448 | PD |
| 3822 | 419271 | 382366 | PD |
| 3822 | 449035 | 440262 | PD |
| 3823 | 419272 | 395585 | PD |

| | | | |
|---|---|---|---|
| 3823 | 449036 | 440267 | PD |
| 3824 | 419579 | 395592 | PD |
| 3824 | 468279 | 449614 | PD |
| 3825 | 419273 | 393639 | PD |
| 3825 | 504450 | 549048 | PD |
| 3826 | 419274 | 395600 | PD |
| 3826 | 468280 | 449625 | PD |
| 3828 | 419580 | 395605 | PD |
| 3828 | 468281 | 449661 | PD |
| 3829 | 419581 | 395614 | PD |
| 3830 | 419582 | 412202 | PD |
| 3830 | 495006 | 468929 | PD |
| 3831 | 419583 | 402267 | PD |
| 3832 | 419584 | 412209 | PD |
| 3832 | 495007 | 468935 | PD |
| 3834 | 419585 | 415724 | PD |
| 3834 | 504454 | 473094 | PD |
| 3835 | 436875 | 415731 | PD |
| 3838 | 436075 | 423748 | PD |
| 3838 | 504456 | 515249 | PD |
| 3869 | 436077 | 415744 | PD |
| 3870 | 363956 | 395313 | PD |
| 3870 | 486557 | 415751 | PD |
| 4005 | 419890 | 397646 | PD |
| 4011 | 418504 | 402285 | PD |
| 4019 | 418710 | 362640 | PD |
| 4019 | 446143 | 417057 | PD |
| 4021 | 419277 | 430178 | PD |
| 4022 | 418712 | 365294 | PD |
| 4022 | 446145 | 417065 | PD |
| 4029 | 468288 | 468943 | PD |
| 4030 | 363959 | 356036 | PD |
| 4030 | 419596 | 415756 | PD |
| 4030 | 495322 | 468949 | PD |
| 4034 | 388585 | 367425 | PD |
| 4034 | 436083 | 423755 | PD |
| 4035 | 388587 | 369183 | PD |
| 4035 | 436084 | 423762 | PD |
| 4035 | 504466 | 475680 | PD |
| 4038 | 388590 | 367446 | PD |
| 4038 | 436085 | 430210 | PD |
